# Association of Adverse Prenatal Exposure Burden with Persistent Psychopathology and Accelerated Cortical Thinning in Youth

**DOI:** 10.1101/2025.07.23.25331227

**Authors:** Dongmei Zhi, Sofia A. Perdomo, Liam R. Arteaga, Dylan E. Hughes, Erin C. Dunn, Phil H. Lee, A. Eden Evins, Harrison T. Reeder, Scott E. Hadland, Alysa E. Doyle, Jacqueline A. Clauss, Jing Sui, Joshua L. Roffman, Jodi M. Gilman

## Abstract

**Importance:** Adverse prenatal exposures (APEs) often co-occur and independently associate with risk for childhood psychopathology. Whether exposure to multiple APEs associates with persistent clinical effects through adolescence or underlying changes in brain maturation remains uncertain.

**Objective:** To evaluate longitudinal associations among cumulative APE burden, risk for psychopathology, and age-related cortical thinning in adolescents.

**Design, Setting, and Participants:** This cohort study analyzed 4-year follow-up data from the Adolescent Brain Cognitive Development (ABCD) Study, which enrolled 11,868 youth aged 9 to 10 years beginning in 2016. Sibling-comparison analysis was performed on 414 non-adopted sibling pairs with discordant APEs. Statistical analysis occurred from March to June 2025.

**Exposures:** Cumulative APE burden was calculated by summing six binary prenatal exposures that independently associated with psychopathology at baseline: unplanned pregnancy; early maternal prenatal alcohol, tobacco, or marijuana use; complicated pregnancy; and complicated birth.

**Main Outcomes and Measures:** Outcomes included annual Child Behavior Checklist (CBCL) scores of dimensional psychopathology, using both continuous and thresholded outcomes; and biennial cortical thickness measures from structural magnetic resonance imaging, analyzed using linear mixed-effects models.

**Results:** Of 8,515 singleton children (4,055 females [47.6%]), 78% were exposed to at least one APE. Multiple APEs persistently and dose-dependently associated with increased odds of clinically significant psychopathology (CBCL total problems: odds ratio=2.01-6.75; corrected P=.0065-1.31×10^-13^). Associations of APEs with attention-deficit/hyperactivity disorder symptoms attenuated over time (interaction: F=13.51; corrected P=7.13×10^-8^), while those with depressive symptoms potentiated (interaction: F=5.82; corrected P=.0019). Greater APE burden associated with accelerated age-related cortical thinning in 36 of 68 cortical regions (interactions: F=3.26–8.89; corrected P’s=0.039-4.86×10^-4^). Siblings with more exposures demonstrated persistently higher CBCL total problems (T=2.25; P=0.025) and accelerated cortical thinning (interactions: T=-3.00–-2.10; P’s<.05) in 5 of the 36 regions implicated in the larger sample.

**Conclusions and Relevance:** Multiple prenatal adversities associated with altered developmental trajectories of psychopathology and cortical maturation into mid-adolescence. These findings highlight the importance of fetal programming to mental health across life course, and the need for additional study of risk and resiliency-conferring factors in utero.

**Key Points:** *Question:* Is adverse prenatal exposure (APE) burden associated with long-term changes in psychopathology and underlying cortical maturation trajectories in children?

*Findings:* In this 4-year follow-up cohort study of 8,515 singleton children in the ABCD Study, exposure to multiple APEs associated with persistent, dose-dependent increases in the risk of clinically significant psychopathology and diffuse acceleration in age-associated cortical thinning during adolescence. Findings were further supported by within-family comparisons in 414 sibling pairs discordant for APEs.

*Meaning:* These findings associate multiple prenatal adversities with enduring and developmentally dynamic effects on both cortical development and mental health in adolescence. The results emphasize the need for early identification, longitudinal monitoring, and neurodevelopmentally informed prevention strategies for at-risk youth.

## INTRODUCTION

Converging evidence from epidemiologic,^1^ genomic,^2^ neuroimaging,^3^ and other translational neuroscience studies^4,5^ implicates fetal brain development as a sensitive period for risk of subsequent psychopathology, in both childhood and adulthood.^1,6^ Adverse prenatal exposures (APEs), such as maternal alcohol,^7,8^ tobacco,^9^ cannabis use,^10,11^ medical complications of pregnancy and childbirth^12,13^ and unplanned pregnancy^14^ may disrupt critical neurodevelopmental processes, thereby laying a biological foundation for subsequent vulnerability to psychopathology.^15^

While much of the literature has focused on the effects of individual APEs on specific psychiatric outcomes,^7,10,16^ prenatal adversities often co-occur,^14^ and may exert cumulative risk effects on neurodevelopment and psychopathology. We previously demonstrated a dose-dependent association between cumulative APE burden and broadly elevated baseline psychopathology in childhood.^14^ However, it remains unclear whether these associations persist or evolve across development, particularly during the transition from childhood to adolescence, a period marked by dynamic changes in psychiatric symptoms.^17^ For example, Attention-Deficit/Hyperactivity Disorder (ADHD) symptoms tend to decline with age,^18^ whereas depressive symptoms tend to increase.^19^ Whether and to what extent exposure to multiple prenatal adversities alters the longitudinal trajectory of psychopathology, potentially increasing vulnerability to psychiatric disorders over time, remains unexplored.

A small but growing literature also links individual APEs to alterations in adolescent brain development, potentially mediating prenatal programming of psychopathology risk. Cortical thickness typically shows a steady age-associated decrease across adolescence,^20^ a pattern thought to reflect synaptic pruning and cortical myelination, providing a clinically relevant development marker.^21^ Exposure to prenatal adversities has been associated with altered cortical thickness in relatively small, cross-sectional studies.^3,22,23^ In turn, deviations in this thinning trajectory—either delayed onset or accelerated decline—have been implicated in heightened risk for psychopathology.^3,24^ However, previous studies are limited by cross-sectional MRI observations and relatively modest sample size, do not control for potential unmeasured confounders, and fail to account for the co-occurrence of multiple APEs. A more holistic, longitudinal account of how multiple APEs shape cortical development throughout early adolescence could clarify underlying mechanisms that may be relevant to diverse forms of emerging psychopathology.

We analyzed data from the ongoing Adolescent Brain Cognitive Development (ABCD) Study^25^ to assess cumulative APE burden on longitudinal trajectories of psychopathology and brain development from age 9 to 15. The ABCD Study is a population-based observational cohort study involving 11,868 U.S. children with serial, harmonized assessments of cortical thickness and dimensional psychopathology. To account for unmeasured familial confounders and shared genetic liability, we leveraged the enrichment of the ABCD Study for multiple family members to conduct similar analyses comparing sibling pairs discordant for APEs.

## METHODS

### Study Design and Participants

The present study analyzed data from the ABCD Study (Release 5.1), which enrolled 11,868 participants aged 9 to 10 years at baseline from 21 U.S. sites. Analyses used data from baseline to 4-year follow-up. Non-adopted singleton pregnancies were included (**eFigure 1**). Detailed cohort characteristics have been reported elsewhere^25^ and are summarized in **Table 1**. Informed consent was obtained from all participants, and the ABCD study was approved by a central institutional review board (IRB) from the University of California, San Diego. This secondary analysis of de-identified ABCD data was exempt from IRB review at Mass General Brigham, Boston, U.S.

**Table 1.** Baseline characteristics for study participants stratified by adverse prenatal exposure burden.

| Characteristic | Participants, No. (%) |  |  |  |  | Statistics | P value |
| --- | --- | --- | --- | --- | --- | --- | --- |
|  | Total<br>(n = 8,515) | No APE<br>(n = 1,871) | One APE<br>(n = 2,945) | Two APEs<br>(n = 2,136) | Three+ APEs<br>(n = 1,563) |  |  |
| Female, No. (%) | 4,055 (47.6%) | 883 (47.2%) | 1,392 (47.3%) | 1,034 (48.4%) | 746 (47.7%) | X<sup>2</sup> = 0.82 | 0.84 |
| Age, y | 9.9 (0.6) | 9.9 (0.6) | 9.9 (0.6) | 9.9 (0.6) | 9.9 (0.6) | F = 0.3 | 0.83 |
| Pubertal stage | 1.9 (0.7) | 1.8 (0.7) | 1.9 (0.7) | 2.0 (0.7) | 2.1 (0.7) | F = 37.76 | 3.43×10<sup>-24</sup> |
| Income-to-needs ratio | 3.6 (2.5) | 4.1 (2.4) | 3.8 (2.6) | 3.5 (2.5) | 2.8 (2.3) | F = 85.61 | 1.81×10<sup>-54</sup> |
| Intracranial volume (mm<sup>3</sup>) | 1,505,490<br>(158,196) | 1,522,239<br>(160,011) | 1,507,354<br>(155,877) | 1,501,502<br>(158,948) | 1,487,278<br>(157,276) | F = 14.15 | 3.40×10<sup>-9</sup> |
| Surface hole number | 28.1 (12.5) | 27.8 (12.1) | 28.1 (12.7) | 28.3 (12.3) | 28.3 (13) | F = 0.59 | 0.62 |
| Scanner |  |  |  |  |  |  |  |
| GE | 4,522 (60.1) | 1,013 (61.1) | 1,550 (59) | 1,145 (60.9) | 814 (60.1) | X<sup>2</sup>=23.09 | 7.67×10<sup>-4</sup> |
| PHILIPS | 758 (10.1) | 194 (11.7) | 285 (10.9) | 177 (9.4) | 102 (7.5) |  |  |
| SIEMENS | 2,216 (29.5) | 446 (26.9) | 782 (29.8) | 555 (29.5) | 433 (32) |  |  |
| Prenatal exposures |  |  |  |  |  |  |  |
| Unplanned pregnancy | 3,379 (39.7%) | 0 | 894 (30.4%) | 1,210 (56.6%) | 1,275 (81.6%) |  |  |
| Early alcohol exposure | 2,157 (25.3%) | 0 | 527 (17.9%) | 668 (31.3%) | 962 (61.5%) |  |  |
| Early tobacco exposure | 1,107 (13%) | 0 | 70 (2.4%) | 261 (12.2%) | 776 (49.6%) |  |  |
| Early marijuana exposure | 501 (5.9%) | 0 | 14 (0.5%) | 56 (2.6%) | 431 (27.6%) |  |  |
| Complicated pregnancy | 3,424 (40.2%) | 0 | 977 (33.2%) | 1,301 (60.9%) | 1,146 (73.3%) |  |  |
| Complicated birth | 2,082 (24.5%) | 0 | 463 (15.7%) | 776 (36.3%) | 843 (53.9%) |  |  |
| Child Behavior Checklist (CBCL) measures at baseline |  |  |  |  |  |  |  |
| Anxious/depressed | 53.6 (6.0) | 52.9 (5.6) | 53.2 (5.7) | 53.7 (6.0) | 54.9 (6.7) | F = 38.33 | 1.34×10<sup>-24a</sup> |
| Withdrawn/depressed | 53.5 (5.8) | 52.7 (4.8) | 53.1 (5.4) | 53.7 (5.9) | 55.1 (6.9) | F = 59.94 | 2.75×10<sup>-38a</sup> |
| Somatic complaints | 55.0 (6.1) | 53.8 (5.2) | 54.5 (5.7) | 55.5 (6.3) | 56.6 (6.9) | F = 67.74 | 3.92×10<sup>-43a</sup> |
| Social problems | 52.8 (4.7) | 51.9 (3.7) | 52.2 (4.0) | 53.1 (4.9) | 54.6 (6.0) | F = 123.03 | 2.23×10<sup>-77a</sup> |
| Thought problems | 53.8 (5.9) | 52.8 (5.0) | 53.2 (5.2) | 54.1 (6.1) | 55.8 (7.1) | F = 90.16 | 3.28×10<sup>-57a</sup> |
| Attention problems | 53.9 (6.1) | 52.7 (4.9) | 53.2 (5.4) | 54.2 (6.1) | 56.2 (7.8) | F = 115.88 | 4.25×10<sup>-73a</sup> |
| Rule-breaking behaviors | 52.7 (4.8) | 51.7 (3.6) | 52.1 (4.0) | 53.0 (5.1) | 54.7 (6.3) | F = 137.16 | 4.93×10<sup>-86a</sup> |
| Aggressive behaviors | 52.8 (5.4) | 51.8 (4.3) | 52.3 (4.7) | 52.9 (5.5) | 54.7 (7.0) | F = 99.03 | 1.11×10<sup>-62a</sup> |
| Internalizing problems | 48.7 (10.6) | 46.6 (10.1) | 47.7 (10.3) | 49.4 (10.5) | 52.0 (10.9) | F = 90.11 | 3.28×10<sup>-57a</sup> |
| Externalizing problems | 45.7 (10.2) | 43.2 (9.3) | 44.6 (9.5) | 46.5 (10.2) | 49.8 (11.2) | F = 142.86 | 2.20×10<sup>-89a</sup> |
| Total problems | 46.0 (11.3) | 42.9 (10.7) | 44.5 (10.8) | 47.1 (11.0) | 50.9 (11.5) | F = 180.08 | 5.63×10<sup>-112a</sup> |
| Depressive symptoms | 53.6 (5.7) | 52.7 (4.8) | 53.3 (5.4) | 53.9 (5.9) | 55.3 (6.7) | F = 66.41 | 2.51×10<sup>-42a</sup> |
| Anxiety symptoms | 53.6 (6.2) | 52.9 (5.5) | 53.1 (5.7) | 53.8 (6.3) | 55.0 (7.1) | F = 43.61 | 6.24×10<sup>-28a</sup> |
| Somatic symptoms | 55.6 (6.7) | 54.4 (5.8) | 55.1 (6.4) | 56.1 (6.9) | 57.2 (7.5) | F = 61.22 | 4.58×10<sup>-39a</sup> |
| ADHD symptoms | 53.2 (5.6) | 52.1 (4.5) | 52.6 (5.0) | 53.5 (5.7) | 55.2 (7.0) | F = 106.91 | 1.47×10<sup>-67a</sup> |
| Oppositional defiant symptoms | 53.5 (5.3) | 52.4 (4.3) | 53.0 (4.8) | 53.7 (5.5) | 55.3 (6.6) | F = 95.02 | 3.27×10<sup>-60a</sup> |
| Conduct symptoms | 52.9 (5.4) | 51.9 (4.2) | 52.2 (4.5) | 53.3 (5.7) | 54.9 (7.0) | F = 120.02 | 1.34×10<sup>-75a</sup> |

Categorical variables were assessed using Chi-square tests, and quantitative measures using analysis of variance (ANOVA) among adverse prenatal exposure (APE) groups. Quantitative measures are presented as means and standard deviations (SD); categorical variables are presented as counts and percentages. Group differences in Child Behavior Checklist (CBCL) T- scores between adverse prenatal exposure (APE) groups are presented. ADHD, Attention- Deficit/Hyperactivity Disorder. <sup>a</sup> Significant after false discovery rate (FDR) correction for 17 multiple comparisons.

### Adverse Prenatal Exposures

Six APEs from the ABCD Developmental History Questionnaire, reported retrospectively by participants’ parents/caregivers at baseline, were included: (1) unplanned pregnancy; (2) early alcohol, (3) tobacco, or (4) marijuana use before knowing of pregnancy; (5) complicated pregnancy; and (6) complicated birth. These APEs were selected based on (1) ≥5% prevalence within the ABCD cohort, and (2) independent association with baseline Child Behavior Checklist (CBCL) total problems, after controlling for numerous demographic and socioeconomic factors from our previous analysis (**eMethods**).^14^ As before,^14^ cumulative APE burden was generated by summing these six binary prenatal exposures, and categorized into four groups for analysis (zero, one, two, and three or more APEs). A full listing of measures and items is provided in **eTable 1**.

### Dimensional Psychopathology

Psychopathology was assessed annually from baseline to 4-year follow-up using the parent/caregiver-reported CBCL.^26^ We analyzed 17 items, comprising 8 sub-syndrome scales; 3 broadband composite scores; and 6 DSM-Oriented symptom scales (**eTable 1**). Higher scores indicate more severe mental health problems, with T scores ≥ 60 for composite scores (≥ 65 for individual symptoms) representing a clinically significant psychopathology.^26^ Raw scores were used in longitudinal analyses, and age- and sex-corrected T scores were used in cross-sectional analyses.

### Cortical Thickness

Imaging acquisition and scanning parameters are described elsewhere.^26–28^ Briefly, Baseline, Year 2, and Year 4 structural MRI scans were acquired using harmonized protocols with Siemens, Philips, or General Electric 3T MRI scanners. Minimally processed T1-weighted images were downloaded from the ABCD Data Archive, and further preprocessed using FreeSurfer version 7.4 (http://surfer.nmr.mgh.harvard.edu/), including removal of non-brain tissue, intensity normalization, and white matter segmentation. The Desikan-Killiany atlas was used to extract cortical thickness of 68 cortical regions. For quality control, we only included scans with a surface hole number (SHN)<63, a validated automated metric that reliably excludes poor-quality images in ABCD with 93% accuracy based on comparison to visually rated images.^29^

## Statistical Analysis

### Association of APE burden with the risk of clinically significant psychopathology

Generalized linear mixed-effects models (GLMMs) with binomial error structure and logit link function were used to estimate associations from APE burden to the odds of clinically significant psychopathology, including 17 binarized CBCL scores (coded as 1 for CBCL composite scores ≥ 60, and other individual measures ≥ 65). Age, sex, pubertal stage (averaged across youth- and parent-reported pubertal development), and income-to-needs ratio (INR, a global measure of socioeconomic status) were included as covariates, and subject ID, family ID, and enrollment site were included as random effects to account for individual, family, and site-level variability.

### Longitudinal association of APE burden with psychopathology and cortical thickness trajectories

Linear mixed-effects models (LMMs) were used to assess associations from APE burden to each continuous CBCL measure and regional cortical thickness, adjusting for the same covariates and random effects as above. For imaging analyses, age^2^, intracranial volume, and SHN were included as additional covariates to account for potential non-linear developmental trajectories of cortical thickness and image quality, with scanner manufacturer as an additional random effect to account for inter-scanner variability.

To further examine whether APE burden alters age-associated trajectories of psychopathology and cortical thickness, each LMM model was then fit again with an additional APE-by-age interaction term. To ensure that findings were not affected by inclusion of lower quality scans, sensitivity analyses were conducted using iteratively higher image quality control thresholds (SHN<37 and SHN<30).^29^

We next tested whether cortical thickness moderated associations between APE burden and age-associated trajectories of CBCL total problems. LMM was constructed with three-way interactions among cortical thickness stratified into tertiles based on baseline cortical thickness, APE burden, and age on CBCL total problems scale. All covariates and random effects listed above were included.

### Sibling-pair validation of discordant APE effects on psychopathology and cortical development

To further validate the primary findings, we conducted secondary, within-family analyses using sibling pairs discordant for APE burden, thereby inherently accounting for unmeasured familial confounders.^29–31^ Within each pair, the sibling with higher exposure was designated as the more-exposed group (coded as 1), and the sibling with lower exposure as the less-exposed group (coded as 0). LMM models used in the main analyses were implemented to assess whether effect sizes of APE burden on CBCL scores and cortical thickness, and their age-associated developmental trajectories remained consistent, adjusting for the same covariates and random effects as above. Because this analysis was constrained to CBCL scores and brain regions that were statistically significant in the full sample, and the sample size in the sibling analysis was considerably smaller (total N=828), we focused primarily on the consistency of effect sizes between the full and sibling samples, and set p<.05 uncorrected for multiple comparisons.

To identify individual APEs potentially driving associations observed in cumulative APE burden models, LMMs were conducted to evaluate associations from individual APEs to CBCL measures and cortical thickness, along with their age-associated trajectories (**eMethods**).

For all analyses, unless otherwise stated, quantitative measures were scaled. Standardized coefficients (β) and associated two-tailed *p*-values from each model were reported for continuous outcomes, and adjusted odds ratios (ORs) for binary variables. Analysis of variance (ANOVA) was used to evaluate overall effects of APE burden on psychopathology and cortical thickness, as well as the overall APE-by-age interactions. False discovery rate (FDR) of 0.05 was used to declare significant findings across 17 CBCL scores and 68 cortical thickness comparisons. Analyses were conducted in R (version 4.4.0) with *lmerTest* package.

## RESULTS

### Characteristics of APE in Children

Of 8,515 singleton children with complete data on the six APEs (4,055 females [47.6%]; mean [SD] age, 9.9 [0.6] years at baseline; **eFigure 1**), 6,644 (78%) had at least one APE (**Figure 1A**), the most prevalent being complicated pregnancy (3,424 [40.2%]) and unplanned pregnancy (3,379 [39.7%]). **Table 1** summarizes baseline characteristics of participants, stratified by cumulative APE burden. No significant differences in age, sex, and SHN were observed between APE groups. Children exposed to more APEs exhibited earlier pubertal development, more severe psychopathology, and greater exposure to socioeconomic disadvantage. High cumulative APE burden was associated with increased psychopathology (**Figure 1B**).

**Figure 1.**
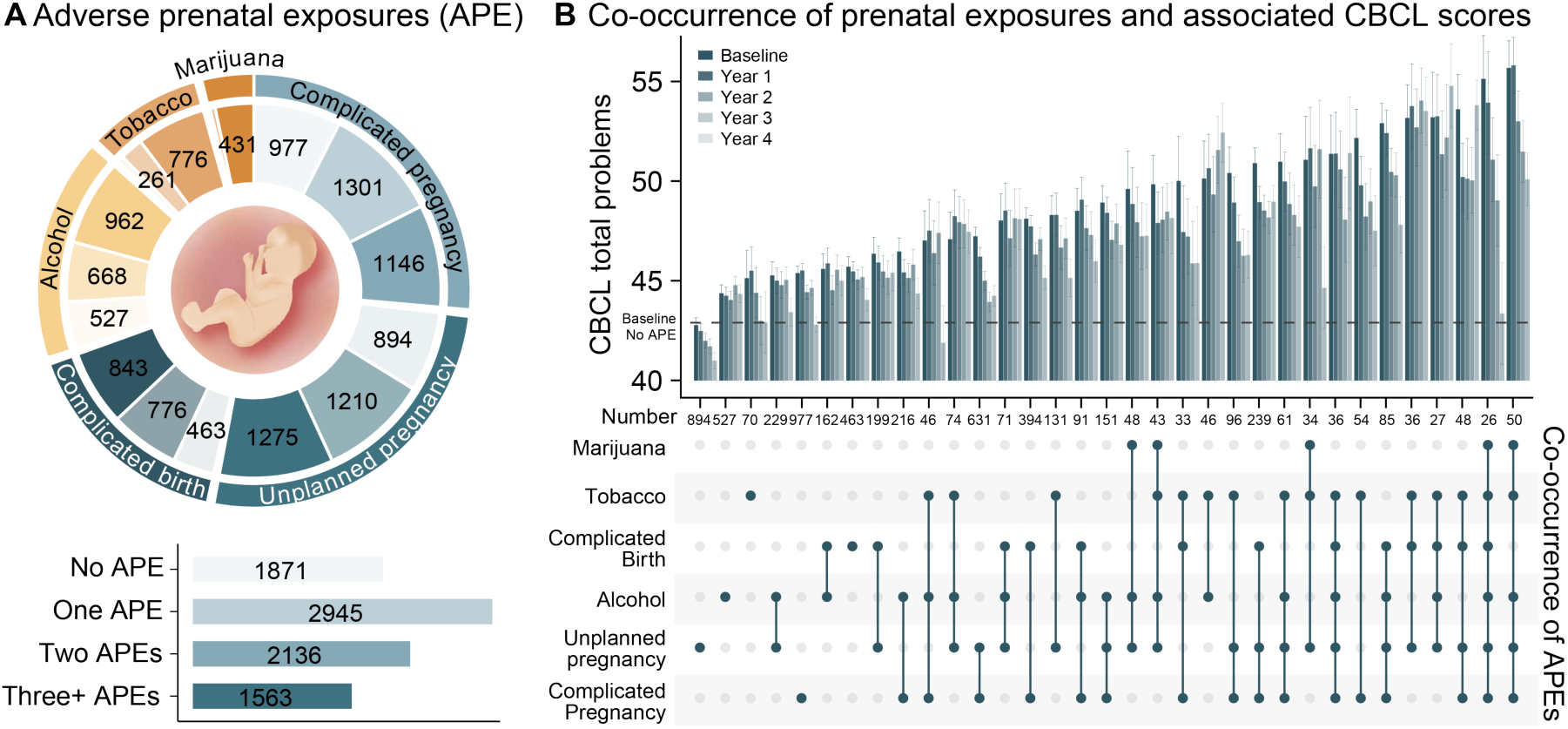
(**A**) Distribution of six adverse prenatal exposures (APEs), including complicated pregnancy, unplanned pregnancy, complicated birth, early alcohol, tobacco, and marijuana use before knowing of pregnancy. Participants were categorized into four groups based on the number of APEs: zero APE, one APE, two APEs, and three or more (three+) APEs. Individual APEs with sample sizes exceeding 100 are labeled within each APE group, where darker color indicates higher APE burden. (**B**) Co-occurrence patterns of APEs (bottom), with co-occurring exposures vertically aligned and connected by dots and lines. The corresponding mean Child Behavior Checklist (CBCL) total problems at Baseline, Year 1, Year 2, Year 3, and Year 4 follow-up are displayed for each exposure combination, ordered by mean baseline CBCL scores, with the number of participants labeled as text. Error bars represent standard errors. The mean baseline CBCL score for the unexposed APE group is indicated by a black dotted line.

### Associations of APE Burden with Clinical Trajectories

From baseline to 4-year follow-up, children with greater APE burden persistently exhibited higher CBCL total problems (**eTable 2**). Although scores declined over time across all APE groups, the stratification by APE burden remained stable (**Figure 2A**). In adjusted GLMM models, cumulative APE burden was associated with clinically significant CBCL total problem scores (χ^2^=69.72; P_FDR_<.001; **Figure 2B**). Specifically, compared with unexposed children, sequentially stronger associations were observed for exposure to one APE (OR, 2.01; 95% CI, 1.28–3.16; P_FDR_=.0065), two APEs (OR, 3.82; 95% CI, 2.39–6.11; P_FDR_<.001), and three or more APEs (OR, 6.75; 95% CI, 4.14–11.02; P_FDR_<.001), demonstrating a dose effect. Results for other binarized CBCL scores are presented in **eFigure 2** and **eTable 3**.

**Figure 2.**
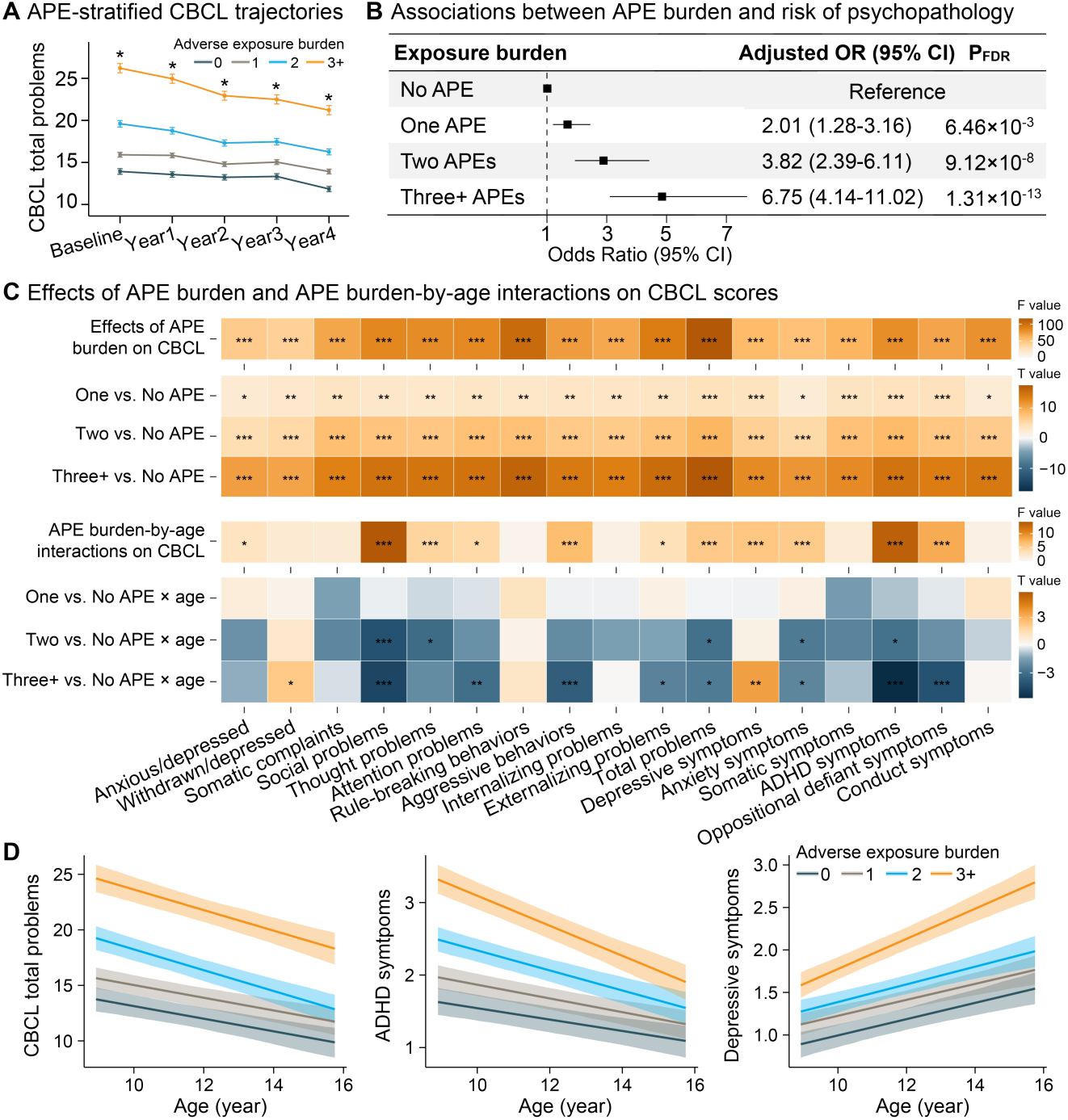
Associations between adverse prenatal exposure (APE) burden and risk of psychopathology and their developmental trajectories, adjusting for age, sex, pubertal stage, socioeconomic status, and random effects for subject ID, family ID, and site effects. (**A**) Longitudinal trajectories of Child Behavior Checklist (CBCL) total problems from baseline to 4-year follow-up, stratified by cumulative number of APEs. From baseline to 4-year follow-up, children exposed to more prenatal adversities persistently exhibited significantly higher CBCL total problems at each time point (*, P_FDR_’s <.001; **eTable 2**), which declined across all APE groups over time, but their stratification by APE burden remained stable. (**B**) Adjusted odds ratios (ORs) and 95% confidence intervals (CIs) for clinically significant psychopathology across cumulative APE levels (one APE, two APEs, three or more [three+] APEs) compared with the unexposed group. The black squares and horizontal lines represent ORs and 95% CIs. (**C**) Effects of APE burden from main effect models (upper panel) and age-by-APE interaction effects from interaction models (lower panel) across APE groups on CBCL scores. Heatmaps illustrate F statistics for overall effects, and T statistics for pairwise comparisons versus the unexposed group. Significant group differences were observed for all CBCL scores across APE groups, with notably altered age-associated trajectories in children exposed to multiple APEs. Asterisks indicate statistical significance (*, P < .05; **, P < .01; ***, P < .001; FDR corrected for 17 comparisons). (**D**) Estimated means (lines) with 95% CIs for age-associated trajectories of CBCL total problems, depressive symptoms, and Attention-Deficit/Hyperactivity Disorder (ADHD) symptoms stratified by APE burden. Note that while stratification of ADHD symptoms by APE burden decreased over time (time x APE interaction F=13.51, P_FDR_=7.13×10^-8^), stratification of depressive symptoms by APE burden increased over time (interaction F=5.82, P_FDR_=.0019), driven in both cases by changes in the most exposed (3+) group.

We next examined associations between APE burden and continuous CBCL scores, as well as APE-by-age interactions on their developmental trajectories. In adjusted LMM models, children with greater APE burden had significantly higher CBCL scores across all domains compared with unexposed children (main effects of APE burden: β=0.06–0.58; T value=2.27–17.63; P_FDR_<0.05; **Figure 2C; eTable 4**). In models with additional APE-by-age interaction terms, significant interactions were observed for 11 of 17 CBCL scores, suggesting that greater APE burden altered age-associated symptom trajectories (**Figure 2C; eTable 5**). For example, greater APE burden was associated with a steeper age-associated decline in CBCL total problems (interaction: F=5.51; P_FDR_=.0023) and ADHD symptoms (interaction: F=13.51; P_FDR_<.001); this effect on ADHD symptoms was particularly pronounced in children exposed to three or more APEs (interaction: β=-0.067; T value=-5.75; P_FDR_<.001), resulting in a progressive narrowing of the symptom gap with unexposed children over time. In contrast, greater APE burden was associated with age-associated increases in DSM-oriented depression symptoms over time (interaction: F=5.82; P_FDR_=.0019), again most evident in three or more APE groups (interaction: β=0.051; T value=3.54; P_FDR_=.0014; **Figure 2D**). Associations between individual APEs and CBCL scores, along with APE-by-age interaction effects, are presented **in eFigure 3, eFigure 4**, **eTable 6**, and **eTable 7**.

### Associations of APE Burden with Altered Cortical Development

One region, the right paracentral cortex, was significantly thicker in children with any APE compared to unexposed children (F=5.96; P_FDR_=.032; **eFigure 5; eTable 8**). In contrast, significant APE-by-age interactions were identified in 36 of 68 cortical regions (β=-0.085–-0.034; F=3.26–8.89; P_FDR_<.05; **Figure 3A; eTable 9**), indicating APE-associated acceleration of cortical thinning across development. Specifically, compared with unexposed children, significant APE-by-age interactions were identified in 0, 19, and 46 cortical regions among children exposed to one, two, and three or more APEs, respectively, indicating a dose-dependent effect of APEs on cortical thinning trajectories. The most prominent effects were observed in right middle temporal cortex (β=-0.078 for three or more APEs; F=8.89; P_FDR_<.001, **Figure 3B**), right inferior parietal cortex (β=-0.085 for three or more APEs; F=6.96; P_FDR_=.0024), and rostral middle frontal cortex (β=-0.078 for three or more APEs; F=6.56; P_FDR_=.0024). To ensure that findings were not affected by inclusion of lower quality scans, this analysis was repeated using iteratively higher image quality control thresholds (SHN<37 and SHN<30).^29^ Main results were largely consistent across different SHN thresholds (**eFigure 6**; **eTable 10**; and **eTable 11**). Associations between individual APEs and cortical thickness, along with APE-by-age interaction effects, are presented in **eFigure 7; eFigure 8; eTable 12**; and **eTable 13**.

**Figure 3.**
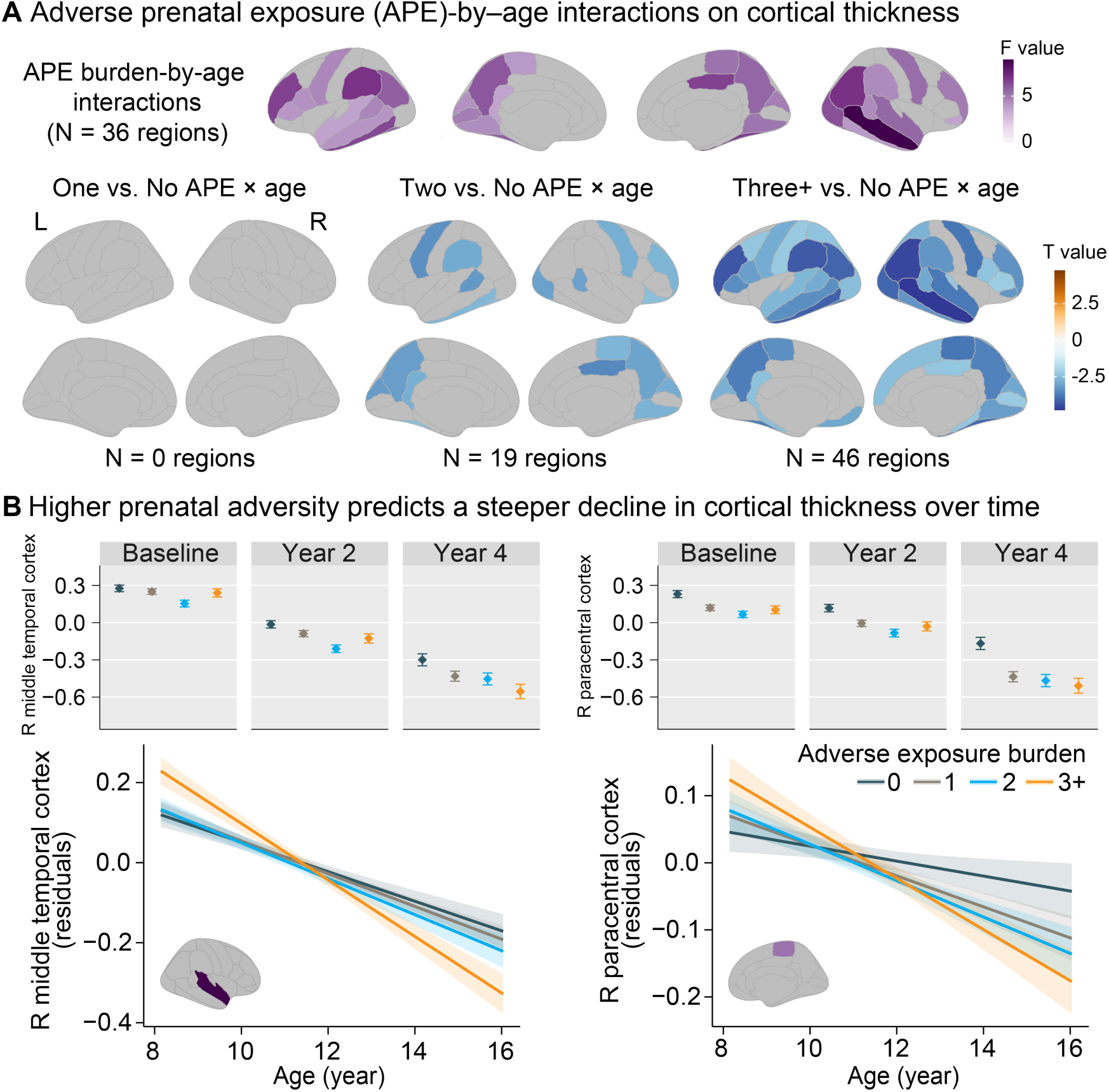
Longitudinal associations of adverse prenatal exposure (APE) burden on cortical thinning. (**A**) Of 68 regions, 36 regions exhibited significant APE-by-age interaction effects on cortical thickness (*, P_FDR_’s <.05, FDR corrected for 68 comparisons). Specifically, compared with unexposed children, significant APE-by-age interactions were identified in 0, 19, and 46 cortical regions among children exposed to one, two, and three or more APEs, respectively. F statistics are shown for overall interaction effects (purple colors), and T statistics are illustrated for pairwise comparison between each APE and no APE group (blue/orange colors). (**B**) Distribution (mean and standard deviation) and aging trajectories for the right middle temporal cortex and right paracentral cortex across APE groups. Higher levels of prenatal adversity were associated with accelerated age-associated cortical thinning. L, left; R, right.

We next examined whether cortical thickness moderated associations between APE group and age-associated trajectories of CBCL total problems by stratifying cortical thickness into tertiles. Of 36 regions examined, nominally significant moderation effects were identified in four regions (**eTable 14**), particularly the left precentral cortex (β=0.13 for three or more APEs at high cortical thickness; F=2.90; uncorrected P=.008). Specifically, children with the thickest cortex and high APE burden exhibited persistently elevated psychopathology symptoms across adolescence (**eFigure 9**). In contrast, children with low or no APE and thinner cortex had more normative age-associated declines in psychopathology.

### Sibling-pair validation of discordant APE effects on psychopathology and cortical development

To validate the primary findings, we conducted within-family analyses using 414 sibling pairs discordant for APE burden (**eTable 15**). More-exposed siblings exhibited higher CBCL total problems across development (β=0.11; T=2.25; P=.025; **Figure 4A; eFigure 10; eTable 16**). As sibling comparisons inherently reduce exposure variability but have limited sample size, we assessed robustness by examining consistency in the direction of interaction effects for three or more APE groups with the strongest interactions in the main analysis. For CBCL scores, 9 of the 11 (82%) significant APE-by-age interactions identified in the main analysis demonstrated consistent directions of interaction effects in CBCL trajectories in sibling comparisons (**Figure 4B; eTable 17**). Specifically, more-exposed siblings had more externalizing problems (β=0.13; T=2.41; P=.017) but the gap diminished over time, whereas an increase in depressive symptoms (β=0.13; T=2.59; P=.010) in the more-exposed siblings became more pronounced over time (**Figure 4C**). Similarly, 34 of 36 (94%) significant interactions on cortical thickness in the main analysis were directionally replicated in sibling comparisons (**Figure 4D; eTable 18**), with five significant regions (β=-0.093–-0.067; T=-2.10–-3.00; P=.0028–.036; **Figure 4E**), including the right middle temporal (β=-0.067; T=-2.10; P=.036) and right paracentral cortices (β=-0.079; T=-2.23; P=.026), where more-exposed siblings showing significantly accelerated cortical thinning.

**Figure 4.**
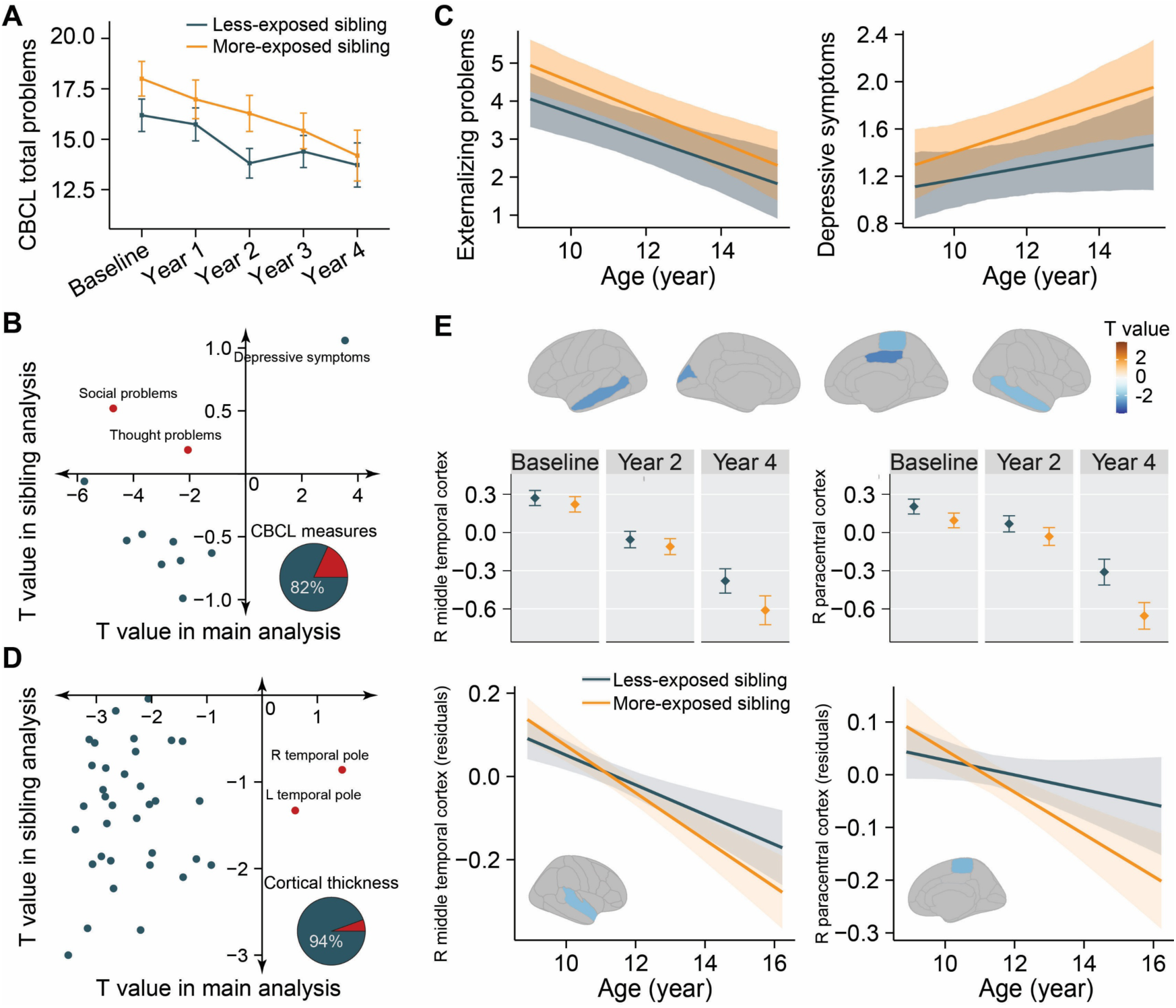
Within-family validation of associations between adverse prenatal exposure (APE), cortical thickness and psychopathology development. (**A**) Longitudinal trajectories of Child Behavior Checklist (CBCL) total problems in 414 matched sibling pairs discordant for APE burden from baseline to 4-year follow-up. More-exposed siblings persistently showed elevated CBCL total problems across development compared with their less-exposed siblings (β=0.11; T = 2.25; P = .025). (**B**) Concordance of APE-by-age interaction effects on CBCL trajectories between the main analysis and sibling comparisons. Among 11 CBCL measures with significant interaction effects for three or more APE groups in the main analysis, 9 (82%) showed consistent direction of interaction effects in the sibling analyses. (**C**) More-exposed siblings had more externalizing problems (β=0.13; T=2.41; P=.017) but the gap diminished over time, whereas an increase in depressive symptoms (β=0.13; T=2.59; P=.010) in the more-exposed siblings became more pronounced over time. (**D**) Concordance of APE-by-age interaction effects on cortical thickness thinning between the main analysis and sibling comparisons. Of 36 cortical regions with significant APE-by-age interactions for three or more APE groups in the main analyses, 34 (94%) showed consistent direction of interaction effects in the within-sibling comparisons. Red dots indicated regions with discordant direction of interaction effects. (**E**) Five significant regions of cortical thickness in observed sibling comparison analyses (β=-0.093–-0.067; T=-2.10–-3.00; P=.0028–.036), all of which were also significant in the full sample. Figures indicate distribution (mean and standard deviation) and age-associated decline of two representative regions, the right middle temporal cortex and right paracentral cortex. More-exposed siblings exhibited accelerated age-associated cortical thinning compared with less-exposed siblings (compare to Figure 3B).

## DISCUSSION

The present study demonstrates that exposure to multiple prenatal adversities associates with a dose-dependent risk for clinically significant psychopathology that persists into mid-adolescence, as well as with diffusely altered trajectories of cortical development. These findings arise from one of the largest available prospective, longitudinal adolescent neurodevelopmental cohort studies; were robust to adjustment for biological, design-related, and socioeconomic factors; and were validated in sibling pairs with discordant APEs, thus accounting for unmeasured familial confounders. The strength of these clinical associations, their persistence over time, and dose-dependence provide convergent support for the importance of fetal life in risk for childhood psychopathology.

While previous studies have primarily focused on isolated prenatal exposures,^7,10,11^ the present findings build upon our previous research^14^ that associated APE burden with dose-dependent increases in psychopathology risk at baseline (age 9-10) in the ABCD Study. The present study provides the first evidence that multiple APEs may shape the developmental trajectories of psychopathology. Specifically, higher APE burden associated with heightened risk for psychopathology in adolescence; however, this effect diminished over time for externalizing problems, whereas intensified for depressive symptoms among children with multiple APEs.

This pattern echoes well-established findings that externalizing disorders (e.g., ADHD) typically emerge in early to mid-childhood, while the incidence of depression rises during adolescence,^32^ highlighting the importance of disorder-specific sensitive periods for assessment, prevention and early intervention.

This study extends previous work associating altered cortical thickness with individual exposures to prenatal alcohol,^23^ tobacco^33^ use, and gestational diabetes.^13^ Although only the paracentral cortex showed a significant main effect of APE burden, 36 regions exhibited significant APE-by-age interactions, highlighting the long-lasting impact of prenatal adversity on cortical maturation. Importantly, these interactions emerged only among children exposed to ≥two APEs, indicating a cumulative impact of prenatal exposures on cortical maturation and emphasizing the critical need for integrating multiple prenatal exposures into a cumulative risk factor.^14^ Furthermore, accelerated cortical thinning was predominantly located in the middle temporal cortex, and rostral middle frontal cortex, implicated in attentional control, memory processing, and visual perception.^34,35^

Additionally, cortical thickness in the precentral cortex may moderate the influence of prenatal exposures on age-related psychopathology trajectories. Notably, individuals with high APE burden and thicker cortex had elevated psychopathology across development, suggesting a potential neurodevelopmental pathway linking prenatal exposures to psychopathology. Further, within-family analyses in sibling pairs with discordant APEs showed consistent results, although limited by small sample size.

Several limitations should be considered when interpreting our findings. First, the ABCD study tended to include families with higher socioeconomic status compared to the US as a whole,^36^ potentially limiting generalizability to populations with socioeconomic adversity. However, findings were robust to the inclusion of income-to-needs ratio, suggesting their broader relevance. Second, retrospective and potentially imprecise self-reporting of prenatal exposures from parents may introduce bias,^14^ such as commonly underreported substance use,^37^ and both over- and underestimated unplanned pregnancy,^38^ which may potentially attenuate associations toward the null hypothesis. Finally, the sample size in the sibling-pair validation was relatively small, limiting statistical power; however, the direction of effects was largely consistent with the primary findings, suggesting their robustness.

In summary, this study demonstrates clear and persistent impacts of multiple APEs on psychopathology and cortical development through mid-adolescence. Leveraging the longitudinal and enriched prenatal exposure data of the ABCD study, we demonstrate that exposure to multiple APEs—particularly three or more—is associated with accelerated cortical thinning and decline in externalizing problems, whereas increase in depressive symptoms, emphasizing the dose-dependent influence of prenatal adversity. These patterns warrant continued follow-up as the ABCD cohort reaches late adolescence, a high-risk period for psychiatric disorders. More broadly, these findings underscore the importance of the prenatal environment to cortical maturation and psychopathology risk, and the need to develop early interventions—perhaps as early as in utero—to mitigate such risk.

## Author Contributions

DZ, SP, DEH, JLR, and JMG have directly accessed and verified the ABCD data reported in the manuscript. JLR and JMG conceptualized the idea and designed the study. DZ, SP, DEH, JLR, and JMG were involved in data curation, formal analysis, investigation, visualization, and validation of the findings. JLR and JMG were involved in funding acquisition, supervision, and project administration. DZ, SP, LA, JLR, and JMG wrote the original draft. All authors were involved in the critical review and editing. All authors approved the final version of the manuscript.

## Data Sharing

The Adolescent Brain Cognitive Development data used in this report are available from the NIMH Data Achieve (https://nda.nih.gov) to Authorized Users.

## Acknowledgments

JLR is supported by R01MH124694. JMG is supported by K02DA052684 and R01DA051540. SEH is supported by R01DA057566 and K18DA059913. The ABCD study is supported by the National Institutes of Health (NIH) and additional federal partners under award numbers U01DA041048, U01DA050989, U01DA051016, U01DA041022, U01DA051018, U01DA051037, U01DA050987, U01DA041174, U01DA041106, U01DA041117, U01DA041028, U01DA041134, U01DA050988, U01DA051039, U01DA041156, U01DA041025, U01DA041120, U01DA051038, U01DA041148, U01DA041093, U01DA041089, U24DA041123, U24DA041147. Additional support for this work was made possible from NIEHS R01-ES032295 and R01-ES031074. A full list of supporters is available online. A list of participating sites and a complete list of the study investigators can be found online. ABCD consortium investigators designed and implemented the study and provided data but did not necessarily participate in analysis or writing of this report. This manuscript reflects the views of the authors and does not reflect the opinions or views of the NIH or ABCD consortium investigators. The ABCD data repository grows and changes over time. The ABCD data used in this report came from http://dx.doi.org/10.15154/z563-zd24.

