## Supplementary files for "Association of Adverse Prenatal Exposure Burden with Persistent Psychopathology and Accelerated Cortical Thinning in Youth"

**eAppendix.** Methods

**eFigure 1.** Flowchart of participant selection

**eFigure 2.** Associations of adverse prenatal exposure (APE) burden with the risk of clinically significant psychopathology

**eFigure 3.** Associations between individual adverse prenatal exposures (APEs) and Child Behavior Checklist (CBCL) scores

**eFigure 4.** Interaction effects of individual adverse prenatal exposures (APEs) and age on CBCL developmental trajectories

**eFigure 5.** Associations between adverse prenatal exposure (APE) burden and cortical thickness

**eFigure 6.** Interactions between adverse prenatal exposure (APE) and age on cortical thinning among children with different surface hole number (SHN) thresholds.

**eFigure 7.** Associations between individual adverse prenatal exposures (APEs) and cortical thickness

**eFigure 8.** Interaction effects of individual adverse prenatal exposures (APEs) and age on cortical thinning

**eFigure 9.** Moderation of the left precentral cortical thickness on the association between APE and age-associated Child Behavior Checklist (CBCL) total problems development

**eFigure 10.** Associations between adverse prenatal exposure (APE) burden and Child Behavior Checklist (CBCL) scores and their developmental trajectories in sibling pairs

**eTable 1.** Summary of demographic and behavioral variables

**eTable 2.** Associations of adverse prenatal exposure (APE) burden with CBCL total problems at each time point

**eTable 3.** Associations of adverse prenatal exposures (APE) burden with the risk of clinically significant psychopathology

**eTable 4.** Associations between adverse prenatal exposure (APE) burden and CBCL scores

**eTable 5.** Interaction effects of adverse prenatal exposure (APE) burden and age on CBCL developmental trajectories

**eTable 6.** Associations between individual adverse prenatal exposure (APE) and Child Behavior Checklist (CBCL) scores

**eTable 7.** Interaction effects of individual adverse prenatal exposure (APE) and age on CBCL developmental trajectories

**eTable 8.** Associations between adverse prenatal exposure (APE) burden and cortical thickness

**eTable 9.** Interaction effects of adverse prenatal exposure (APE) burden and age on developmental trajectories of cortical thickness

**eTable 10.** Interaction effects of adverse prenatal exposure (APE) burden and age on cortical thickness trajectories among children with SHN < 37

**eTable 11.** Interaction effects of adverse prenatal exposure (APE) burden and age on cortical thickness trajectories among children with SHN < 30

**eTable 12.** Associations between individual adverse prenatal exposure (APE) and cortical thickness

**eTable 13.** Interaction effects of individual adverse prenatal exposure (APE) and age on cortical thickness thinning

**eTable 14.** Moderation of cortical thickness on associations between adverse prenatal exposure (APE) and age-associated trajectories of CBCL total problems

**eTable 15.** Baseline characteristics of sibling pairs with discordant adverse prenatal exposures (APEs)

**eTable 16.** Associations of adverse prenatal exposure (APE) group with CBCL scores in sibling pairs

**eTable 17.** Interaction effects of adverse prenatal exposure (APE) and age on CBCL developmental trajectories in sibling pairs

**eTable 18.** Interaction effects of adverse prenatal exposure (APE) group with cortical thickness thinning in sibling pairs

**eAppendix.** Methods

**Study Cohort and Participant**

The present study analyzed data from the Adolescent Brain Cognitive Development (ABCD) Study (Release 5.1), which enrolled 11,868 participants aged 9 to 10 years at baseline from 21 regionally distributed sites across the U.S. Detailed information about sample collection, survey measures, and study protocols has been reported elsewhere.^1^ Participants were excluded if they were adopted, identified as intersex or had unspecified sex, or were non-singleton pregnancies (e.g., twin and triplets) due to known differences in pre- and postnatal life compared with singleton births (**eFigure 1**). Participants with incomplete data on any individual exposures were excluded. Informed consent was obtained from all participants, and the ABCD study was approved by a central institutional review board (IRB) at the University of California, San Diego. This secondary analysis of de-identified ABCD data was exempt from IRB review at Mass General Brigham, Boston, U.S.

**Adverse Prenatal Exposures**

A total of 15 adverse prenatal exposures (APEs) were extracted for each participant and coded as present or absent in our previous study,^2^ including (1) unplanned pregnancy; early maternal use of (2) alcohol, (3) tobacco, (4) marijuana, (5) cocaine, or (6) opiates, before knowing of pregnancy; maternal use of (7) alcohol, (8) tobacco, (9) marijuana, (10) cocaine, or (11) opiates after knowing of pregnancy; (12) caesarian section; (13) complicated pregnancy (coded as present if ≥1 of 13 complications was reported, including severe nausea and vomiting extending past the sixth month or accompanied by weight loss; heavy bleeding requiring bed rest or special treatment; pre-eclampsia, eclampsia, or toxemia; severe gall bladder attack; persistent proteinuria; rubella during first 3 months of pregnancy; severe anemia; UTI; pregnancy-related diabetes; pregnancy-related high blood pressure; previa, abruptio, other problems with placenta; accident or injury requiring medical care; and any other conditions requiring medical care); (14) complicated birth (coded as present if ≥1 of 8 complications occurred, including blue at birth; slow heart beat; did not breathe at first;	convulsions; jaundice needing treatment;	required oxygen;	required blood transfusion; and Rh incompatibility); (15) pre-term birth (coded as present if birth occurred before 37 gestational weeks). Seven APEs were reported in fewer than 5% of participants in the ABCD cohort, and two APEs were not significantly associated with baseline psychopathology, resulting in 6 individual APEs for primary analysis,^2^ including unplanned pregnancy; early alcohol, tobacco, or marijuana exposure; complicated pregnancy; and complicated birth. Cumulative APE burden was generated by summing these six binary prenatal exposures, and categorized into four groups for analysis (zero, one, two, and three or more APEs). A full listing of measures, items, and content is provided in **eTable 1**.

**Exploratory Analysis**

To assess independent associations of individual APEs with CBCL scores, linear mixed-effects models were conducted, simultaneously including all six APEs, with age, sex, pubertal stage, income-to-needs ratio as covariates, and random effects for subject ID, family ID, and site to account for individual, family, and site-level variability.

To further examine the interaction effects between each APE-by-age on CBCL development, LMM model was then fit again with additional all six APE-by-age interaction terms.

Similarly, the same analyses were implemented to investigate associations between six individual APEs and cortical thickness, as well as APE-by-age interactions on cortical thinning. For imaging analyses, age^2^, intracranial volume, and surface hole number (SHN) were included as additional covariates to account for non-linear developmental trajectory of cortical thickness and image quality, and scanner manufacturer was included as an additional random effect to account for inter-scanner variability.

**eFigure 1. Flowchart of participant selection.** Among 11,868 participants enrolled in the ABCD Study, 3 with intersex or unspecified sex, 276 adopted children, 2,153 twins or triplets, and 921 with missing prenatal exposure data were excluded. A total of 8,515 participants were included in the main analysis; of these, 1,514 were also analyzed in the sibling analysis. For quality control of cortical thickness, only imaging scans with surface hole number (SHN) < 63 were included in analyses. Only a subset of participants had completed the Year 4 follow-up assessments in the ongoing ABCD Study (Release 5.1).

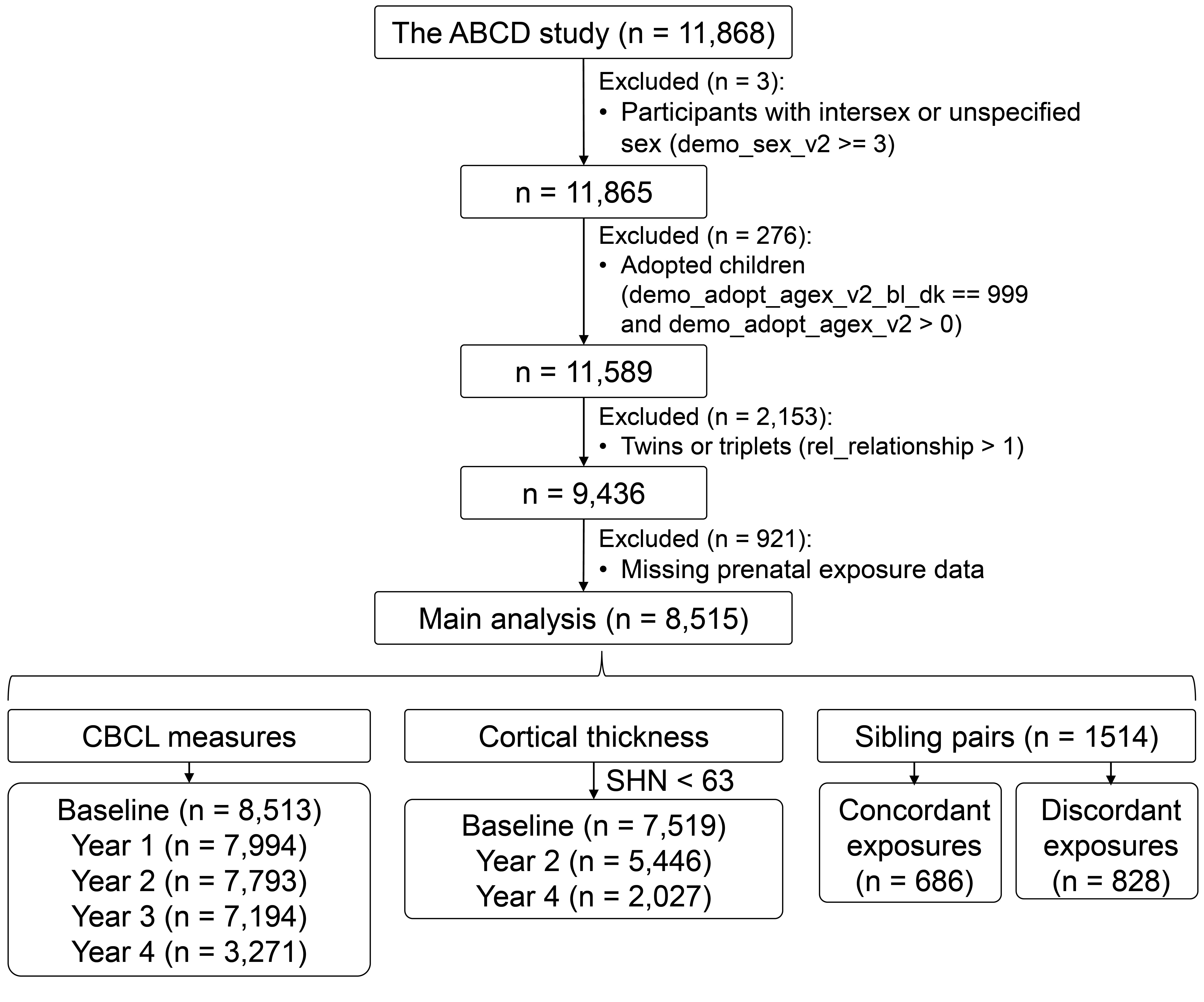

**eFigure 2. Associations of adverse prenatal exposure (APE) burden with the risk of clinically significant psychopathology.** Generalized linear mixed-effects models (GLMMs) with binomial error structure and logit link function were used to estimate associations from APE burden to the odds of clinically significant psychopathology, adjusting for age, sex, pubertal stage, and income-to-needs ratio, and random effects for subject ID, family ID, and enrollment site to account for individual, family, and site-level variability. We illustrate Chi-square statistics for overall effects, and adjusted odds ratios (ORs) for clinically significant psychopathology across APE burden (one APE, two APEs, three or more [three+] APEs) compared with the unexposed group (**eTable 3**). ADHD, attention-deficit/hyperactivity disorder. Asterisks indicate statistical significance (*, P_FDR_ < .05; **, P_FDR_ < .01; ***, P_FDR_ < .001; corrected for 17 multiple comparisons).

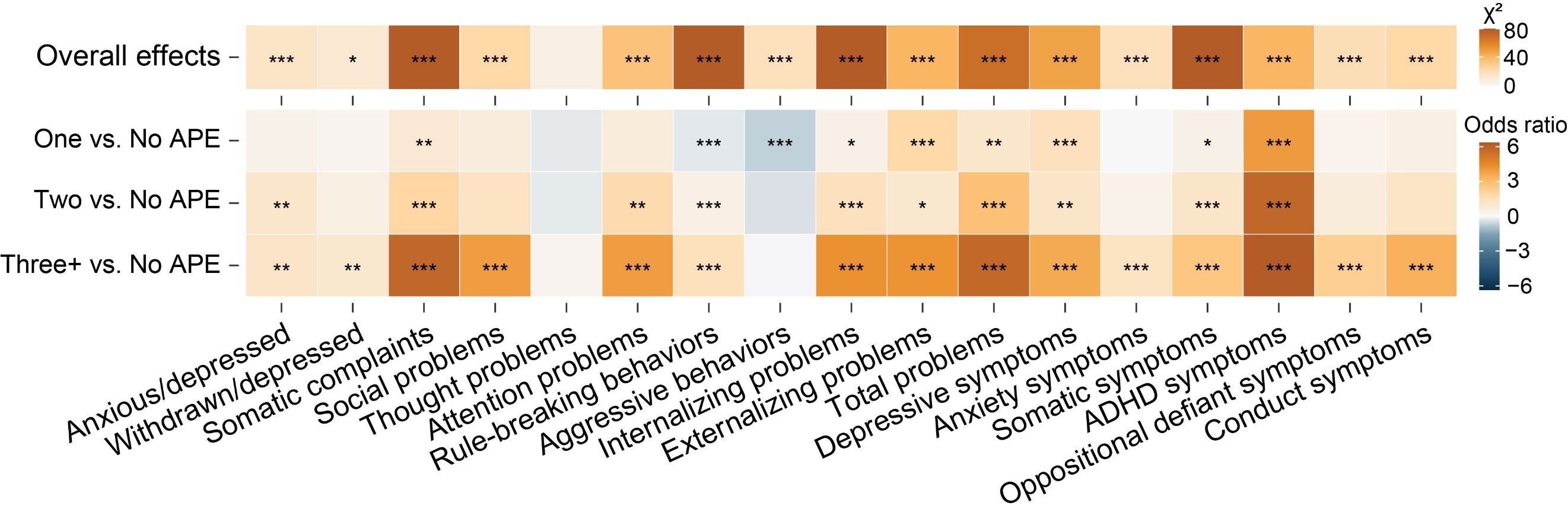

**eFigure 3. Associations between individual adverse prenatal exposures (APEs) and Child Behavior Checklist (CBCL) scores.** Linear mixed-effects models were used to examine associations between six individual APEs and 17 CBCL scores, controlling for age, sex, pubertal stage, socioeconomic status, and random effects for subject ID, family ID, and site effects. Within each model, all six APEs were entered simultaneously to determine the independent effects of each exposures on each CBCL score. Heatmaps illustrate T statistics for associations between six APEs and CBCL scores. Complicated birth, complicated pregnancy, tobacco and alcohol use were significantly associated with all CBCL measures. Asterisks indicate statistical significance (*, P_FDR_ < .05; **, P_FDR_ < .01; ***, P_FDR_ < .001; FDR corrected for 17 multiple comparisons; **eTable 6**).

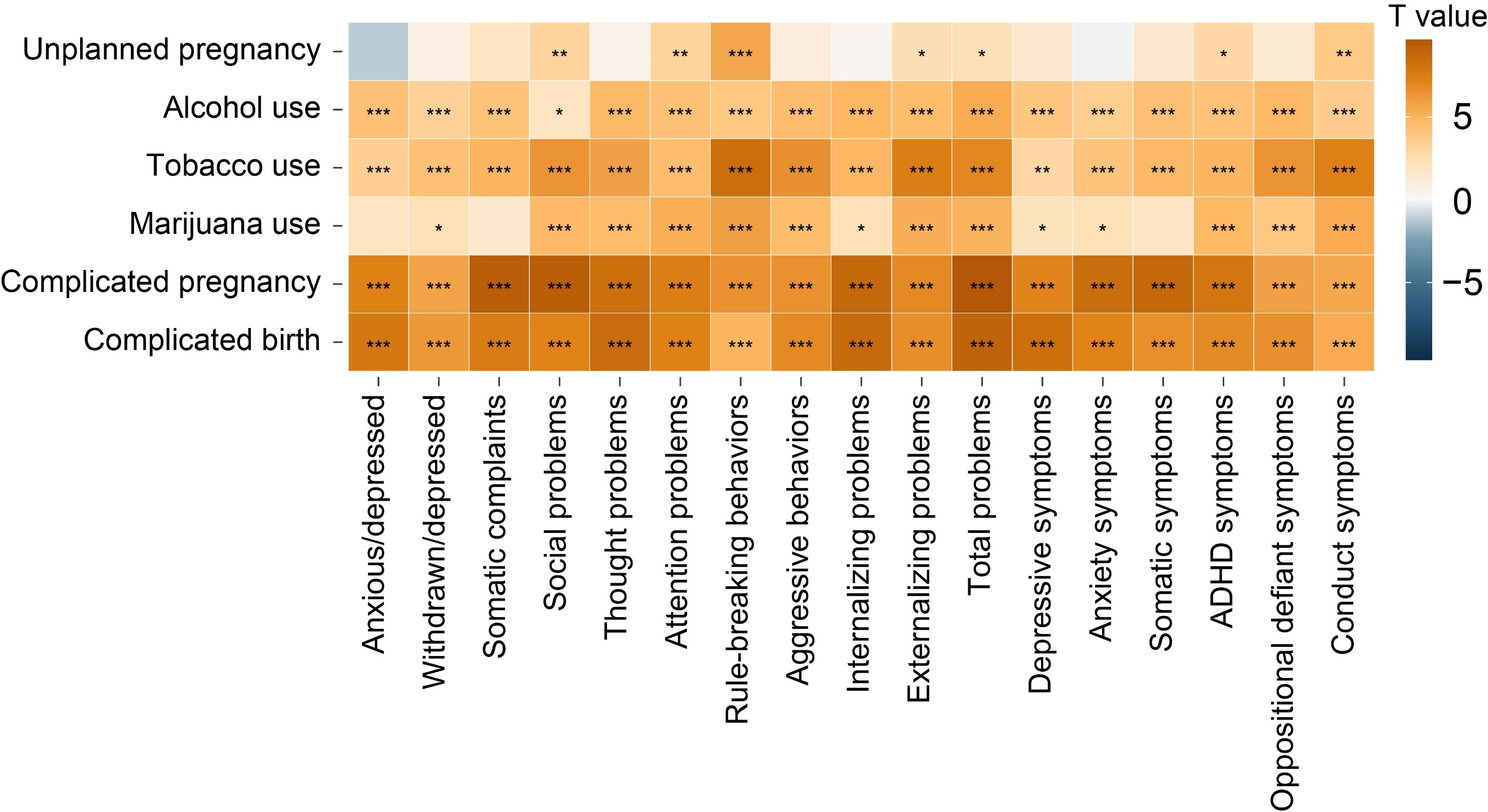

**eFigure 4. Interaction effects of individual adverse prenatal exposures (APEs) and age on CBCL developmental trajectories.** To examine APE-by-age interactions on CBCL developmental trajectories, linear mixed-effects models were fit again, where CBCL scores were modeled as continuous dependent variables, with six individual APEs, age, and six APE-by-age interaction terms as fixed effects, adjusting for sex, pubertal stage, socioeconomic status, and random effects for subject ID, family ID, and site effects. Within each model, all six APEs and six APE-by-age interactions were entered simultaneously to determine independent effects of each exposures on each CBCL score. Standardized beta coefficients and standard errors (SE) were reported for interaction effects between age and individual APEs (*, P_FDR_ < .05; **, P_FDR_ < .01; ***, P_FDR_ < .001; corrected for 17 multiple comparisons; **eTable 7**).

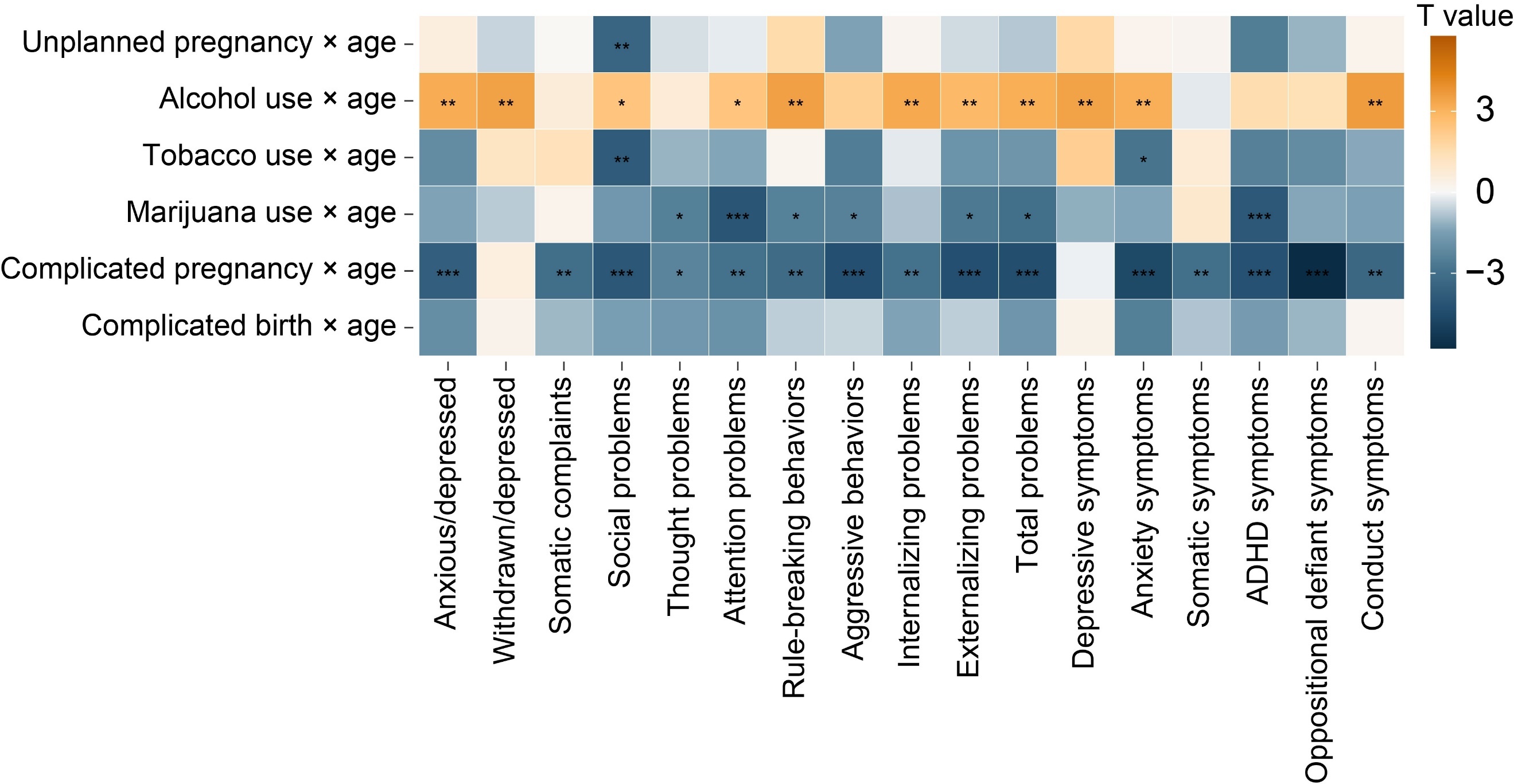

**eFigure 5. Associations between adverse prenatal exposure (APE) burden and cortical thickness.** We examined associations between APE burden and cortical thickness across 68 regions using linear mixed-effects models, controlling for age, age^2^, sex, pubertal stage, socioeconomic status, intracranial volume, surface hole number, and random effects of subject ID, family ID, scanner, and site. T statistics for group differences between each APE group (one APE, two APEs, three or more [three+] APEs) compared with the unexposed group, and F statistics for overall effects were illustrated (P_FDR_ < .05, corrected for 68 multiple comparisons; **eTable 8**).

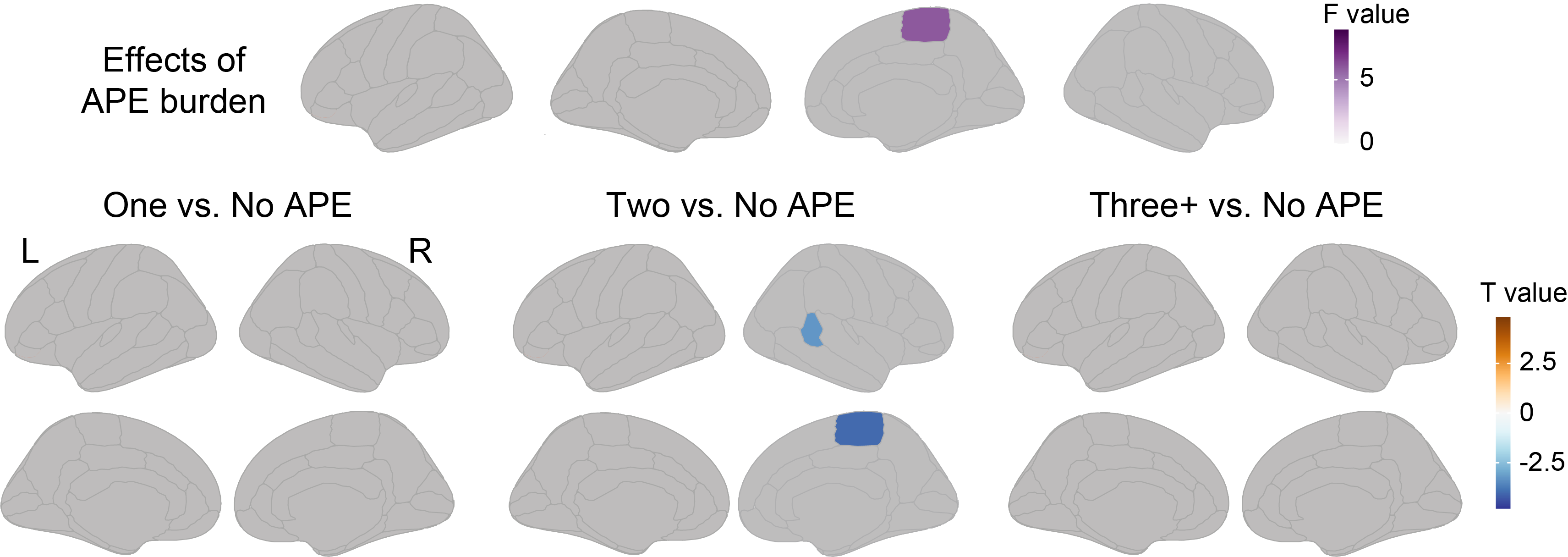

**eFigure 6**. **Interactions between adverse prenatal exposure (APE) and age on cortical thinning among children with different surface hole number (SHN) thresholds.** (**A**) Among children with SHN < 63 (N = 6544), 36 of 68 regions exhibited significant APE-by-age interactions on cortical thinning. Compared with unexposed children, significant interactions were observed in 0, 19, and 46 cortical regions among those exposed to one, two, and three or more APEs, respectively. (**B**) Among children with SHN < 37 (N = 5939), 37 of 68 regions exhibited significant APE-by-age interactions on cortical thickness. Compared with unexposed children, significant interactions were observed in 0, 19, and 42 cortical regions among those exposed to one, two, and three or more APEs, respectively (**eTable 10**). (**C**) Among children with SHN < 30 (N = 5444), 24 of 68 regions exhibited significant APE-by-age interactions on cortical thickness. Compared with unexposed children, significant interactions were observed in 0, 16, and 37 cortical regions among those exposed to one, two, and three or more APEs, respectively (**eTable 11**). F statistics are shown for overall interaction effects (purple colors), and T statistics are illustrated for pairwise comparison between each APE and no APE group (blue/orange colors).

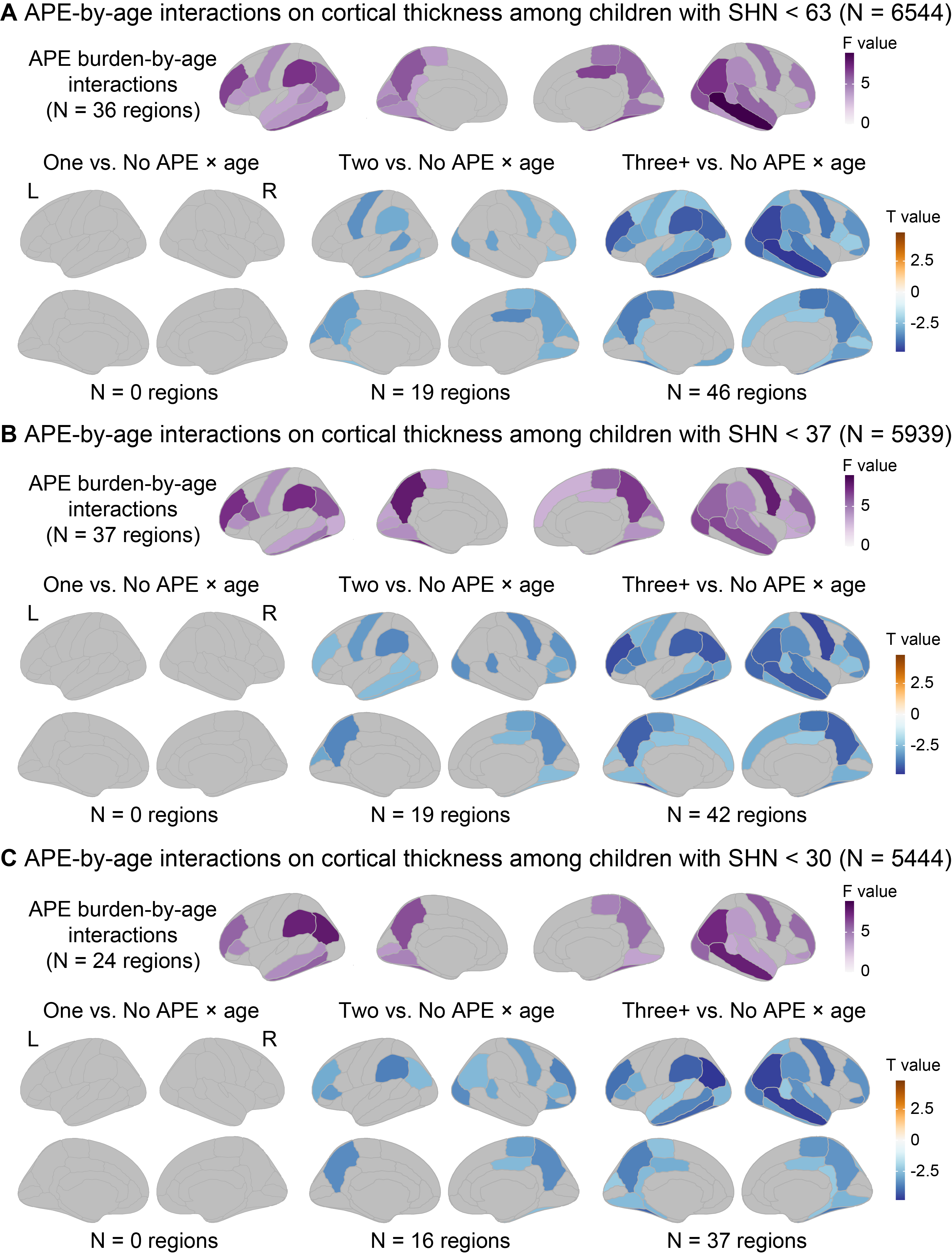

**eFigure 7. Associations between individual adverse prenatal exposures (APEs) and cortical thickness.** Linear mixed-effects models were used to examine associations between six individual APEs and cortical thickness across 68 regions, adjusting for age, age^2^, sex, pubertal stage, intracranial volume, surface hole number, socioeconomic status, and random effects for subject ID, family ID, scanner, and site effects. Within each model, all six APEs were entered simultaneously to determine the independent effects on cortical thickness. T statistics for associations between six APEs and cortical thickness were illustrated with uncorrected P < .05 (left) and FDR corrected P < .05 for 68 multiple comparisons (right; **eTable 12**).

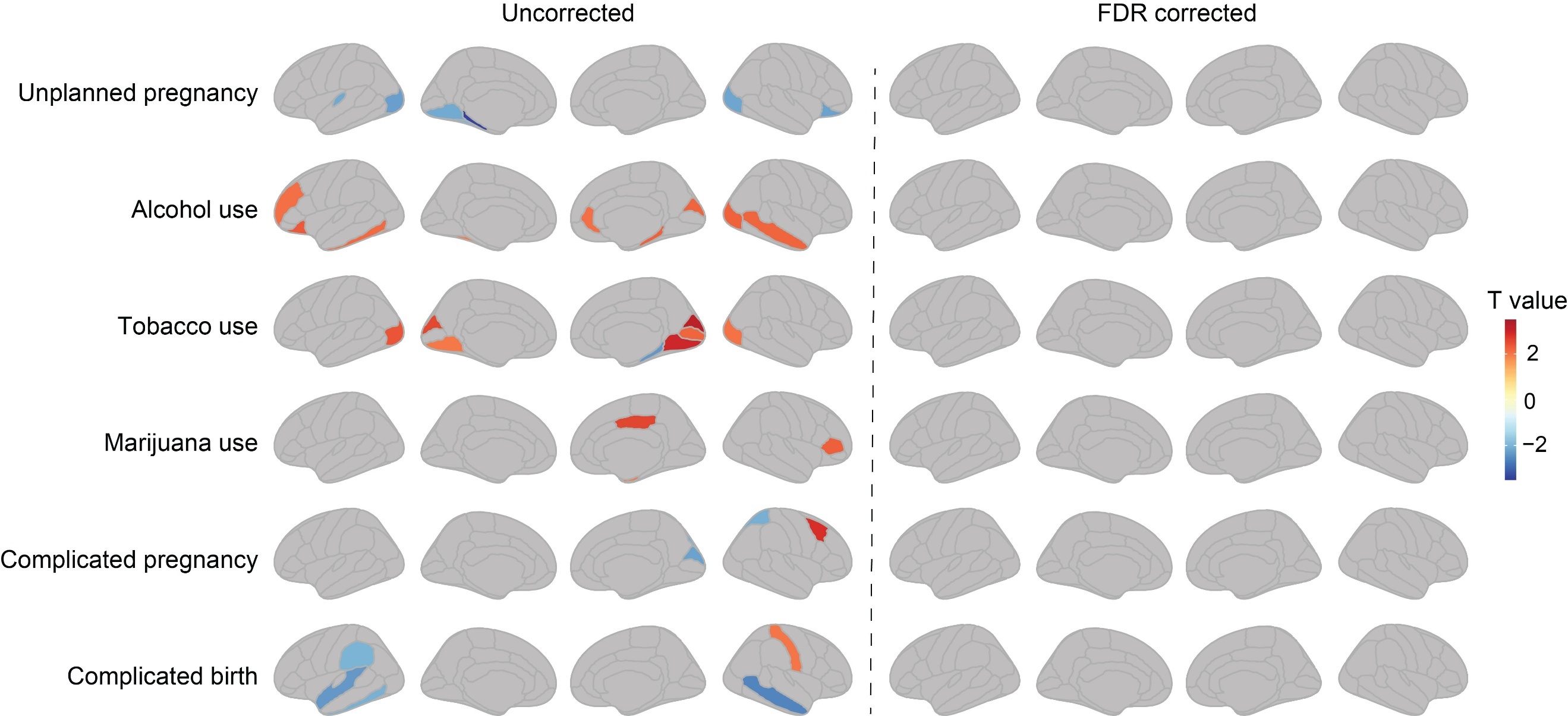

**eFigure 8. Interaction effects of individual adverse prenatal exposures (APEs) and age on cortical thinning.** To examine APE-by-age interactions on cortical thinning, linear mixed-effects models were conducted, where cortical thickness were modeled as continuous dependent variables, with six individual APEs, age, and six APE-by-age interaction terms as fixed effects, adjusting for age, age^2^, sex, pubertal stage, intracranial volume, surface hole number, socioeconomic status, and random effects for scanner, subject ID, family ID, and site. Within each model, all six APEs and six APE-by-age interactions were entered simultaneously to determine independent effects on cortical thickness. T statistics for associations between six APE-by-age interactions on cortical thickness were illustrated with uncorrected P < .05 (left) and FDR corrected P < .05 for 68 multiple comparisons (right; **eTable 13**).

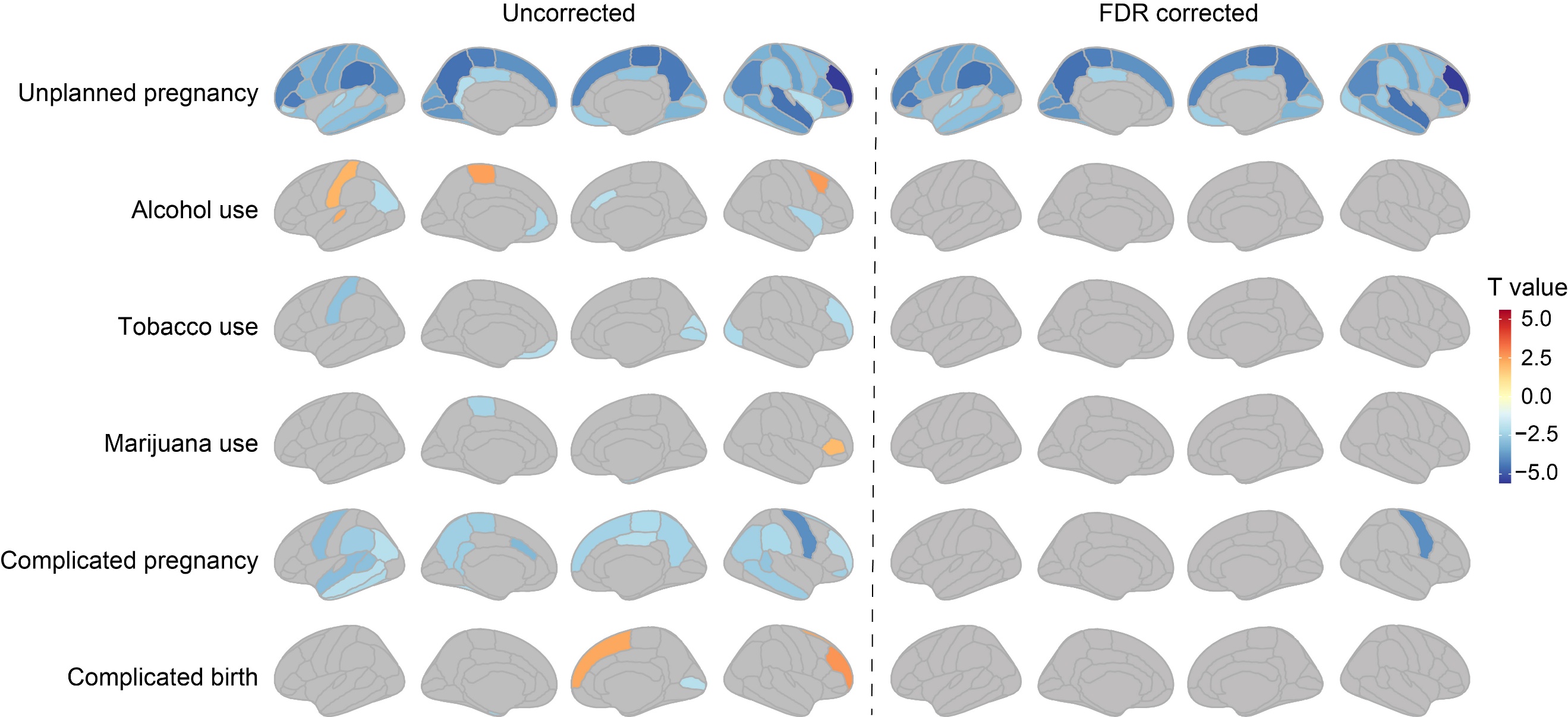

**eFigure 9. Moderation of the left precentral cortical thickness on the association between APE and age-associated Child Behavior Checklist (CBCL) total problems development.** Age-associated trajectories of CBCL total problems are shown across tertiles of baseline cortical thickness (low, medium, high). Children with thicker left precentral cortex and more APEs had elevated psychopathology over time, suggesting that cortical thickness moderated age-associated changes in psychopathology across cumulative APE burden. The shaded areas around the lines denote 95% confidence intervals.

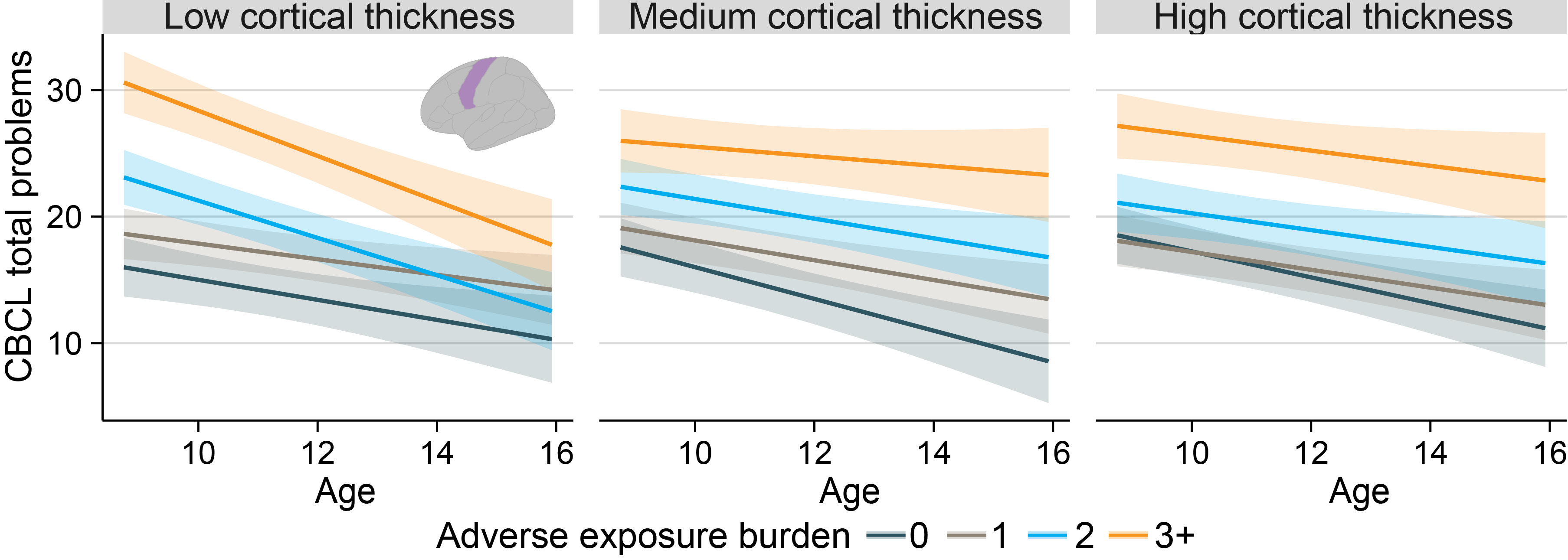

**eFigure 10. Associations between adverse prenatal exposure (APE) burden and Child Behavior Checklist (CBCL) scores and their developmental trajectories in sibling pairs.** Group differences in 17 CBCL measures between more- and less-exposed siblings were examined using linear mixed-effects models, controlling for age, sex, pubertal stage, socioeconomic status, and random effects for subject ID, family ID, and site effects. Heatmaps illustrate T statistics for group differences (top) and age-by-group interaction effects (bottom) between more- and less-exposed groups on CBCL measures. Six nominally significant group differences were identified, with the more-exposed group exhibiting higher rule-breaking, aggressive, externalizing, and total problems, as well as conduct and depressive symptoms. Asterisks indicate statistical significance with uncorrected P value < .05.

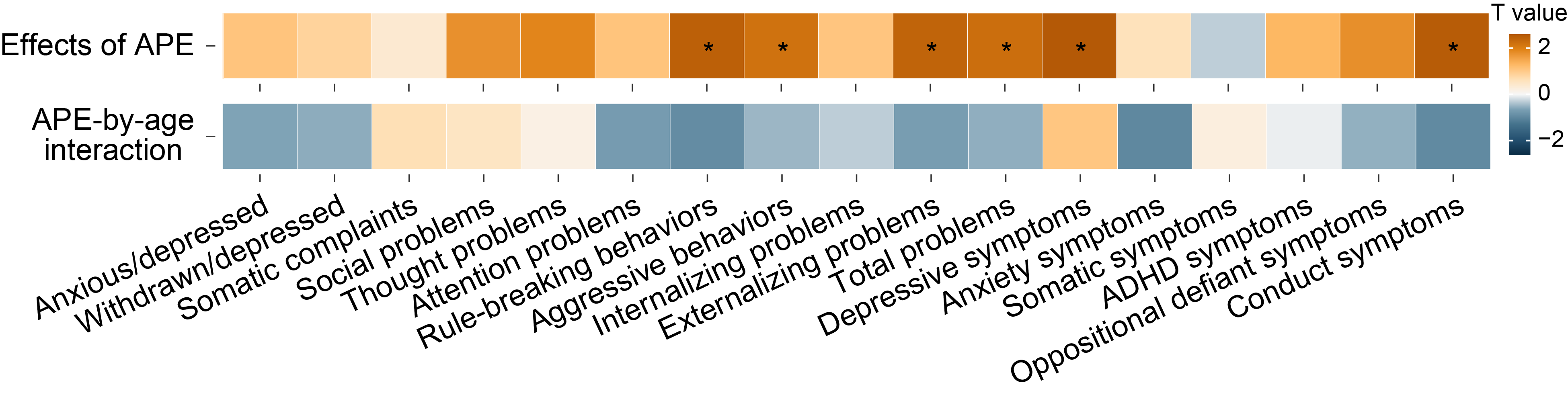

**eTable 1. Summary of demographic and behavioral variables**

| **Measures** | **Description** | **Variable name in ABCD 5.1 website** | **NDA_Short_Name** |
| --- | --- | --- | --- |
| **Sex** | Male; Female. | demo_sex_v2 | abcd_p_demo |
| **Age** | Age in months at the time of the interview | [interview_age](https://nda.nih.gov/general-query.html?q=query=data-element%20~and~%20searchTerm=interview_age%20~and~%20orderBy=elementName%20~and~%20orderDirection=Ascending) | abcd_y_lt |
| **Pubertal stage** | Averaged pubertal development reported by parent and youth | pds_p_ss_female_category_2;  pds_p_ss_male_category_2;  pds_y_ss_female_category_2;  pds_y_ss_male_cat_2. | ph_p_pds |
| **Income-to-needs ratio^a^** | Household income level;  Number of people in the household. | demo_comb_income_v2_l;  demo_roster_v2_l. | abcd_p_demo |
| **Family ID** | Participants belonging to the same family share a family ID | rel_family_id | abcd_p_demo |
| **Site** | Site ID at each event | site_id_l | abcd_y_lt |
| **Prenatal exposures** |  |  |  |
| Unplanned pregnancy | Was your pregnancy with this child a planned pregnancy?  No = 1; Yes = 0. | devhx_6_p | ph_p_dhx |
| Early alcohol exposure | Before knowing of pregnancy. Alcohol?  Yes = 1; No = 0. | devhx_8_alcohol |  |
| Early tobacco exposure | Before knowing of pregnancy. Tobacco?  Yes = 1; No = 0. | devhx_8_tobacco |  |
| Early marijuana exposure | Before knowing of pregnancy. Marijuana?  Yes = 1; No = 0. | devhx_8_marijuana |  |
| Complicated pregnancy | During the pregnancy with this child, did you/biological mother have any of the following conditions? (1) Severe nausea and vomiting extending past the 6th month or accompanied by weight loss; (2) Heavy bleeding requiring bed rest or special treatment; (3) Pre-eclampsia, eclampsia, or toxemia; (4) Severe gall bladder attack; (5) Persistent proteinuria; (6) Rubella during first 3 months of pregnancy; (7) Severe anemia; (8) Urinary tract infections; (9) Pregnancy-related diabetes; (10) Pregnancy-related high blood pressure ; (11) Previa, abruptio, other problems with placenta; (12) An accident or injury requiring medical care; (13) Any other conditions requiring medical care.  At least one of the above items was endorsed = 1; otherwise, No = 0. | devhx_10a3_p;  devhx_10b3_p; devhx_10c3_p; devhx_10d3_p; devhx_10e3_p;  devhx_10f3_p;  devhx_10g3_p; devhx_10h3_p;  devhx_10i3_p;  devhx_10j3_p;  devhx_10k3_p;  devhx_10l3_p;  devhx_10m3_p. |  |
| Complicated birth | Did he/she have any of the following complications at birth? (1) Blue at birth;  (2) Slow heartbeat; (3) Did not breathe at first; (4) Convulsions; (5) Jaundice needing treatment; (6) Required oxygen; (7) Required blood transfusion; (8) Rh incompatibility.  At least one of the above items was endorsed = 1; otherwise, No = 0. | devhx_14a3_p; devhx_14b3_p; devhx_14c3_p; devhx_14d3_p; devhx_14e3_p; devhx_14f3_p; devhx_14g3_p; devhx_14h3_p. |  |
| **Child Behavior Checklist (CBCL) measures** | |  |  |
| Anxious/depressed | AnxDep CBCL Syndrome Scale (raw score/t-score) | cbcl_scr_syn_anxdep_r;  cbcl_scr_syn_anxdep_t. | mh_p_cbcl |
| Withdrawn/depressed | WithDep CBCL Syndrome Scale (raw score/t-score) | cbcl_scr_syn_withdep_r;  cbcl_scr_syn_withdep_t. |  |
| Somatic complaints | Somatic CBCL Syndrome Scale (raw score/t-score) | cbcl_scr_syn_somatic_r;  cbcl_scr_syn_somatic_t. |  |
| Social problems | Social CBCL Syndrome Scale (raw score/t-score) | cbcl_scr_syn_social_r;  cbcl_scr_syn_social_t. |  |
| Thought problems | Thought CBCL Syndrome Scale (raw score/t-score) | cbcl_scr_syn_thought_r;  cbcl_scr_syn_thought_t. |  |
| Attention problems | Attention CBCL Syndrome Scale (raw score/t-score) | cbcl_scr_syn_attention_r;  cbcl_scr_syn_attention_t. |  |
| Rule-breaking behaviors | RuleBreak CBCL Syndrome Scale (raw score/t-score) | cbcl_scr_syn_rulebreak_r;  cbcl_scr_syn_rulebreak_t. |  |
| Aggressive behaviors | Aggressive CBCL Syndrome Scale (raw score/t-score) | cbcl_scr_syn_aggressive_r;  cbcl_scr_syn_aggressive_t. |  |
| Internalizing problems | Internal CBCL Syndrome Scale (raw score/t-score) | cbcl_scr_syn_internal_r;  cbcl_scr_syn_internal_t. |  |
| Externalizing problems | External CBCL Syndrome Scale (raw score/t-score) | cbcl_scr_syn_external_r;  cbcl_scr_syn_external_t. |  |
| Total problems | TotProb CBCL Syndrome Scale (raw score/t-score) | cbcl_scr_syn_totprob_r;  cbcl_scr_syn_totprob_t. |  |
| Depressive symptoms | Depress CBCL DSM5 Scale  (raw score/t-score) | cbcl_scr_dsm5_depress_r;  cbcl_scr_dsm5_depress_t. |  |
| Anxiety symptoms | AnxDisord CBCL DSM5 Scale (raw score/t-score) | cbcl_scr_dsm5_anxdisord_r;  cbcl_scr_dsm5_anxdisord_t. |  |
| Somatic symptoms | SomaticPr CBCL DSM5 Scale (raw score/t-score) | cbcl_scr_dsm5_somaticpr_r;  cbcl_scr_dsm5_somaticpr_t. |  |
| ADHD symptoms | ADHD CBCL DSM5 Scale  (raw score/t-score) | cbcl_scr_dsm5_adhd_r;  cbcl_scr_dsm5_adhd_t. |  |
| Oppositional defiant symptoms | Opposit CBCL DSM5 Scale  (raw score/t-score) | cbcl_scr_dsm5_opposit_r;  cbcl_scr_dsm5_opposit_t. |  |
| Conduct symptoms | Conduct CBCL DSM5 Scale  (raw score/t-score) | cbcl_scr_dsm5_conduct_r;  cbcl_scr_dsm5_conduct_t. |  |

Abbreviations: ADHD, attention-deficit/hyperactivity disorder. ^a^: Income-to-needs ratio was calculated as a mid-point of total combined family income categories (~$5,000; $5,000 ~ $11,999; $12,000 ~ $15,999; $16,000 ~ $24,999; $25,000 ~ $34,999; $35,000 ~ $49,999; $50,000 ~ $74,999; $75,000 ~ $99,999; $100,000 ~ $199,999; $200,000~) divided by 2017 federal poverty guideline according to household sizes (https://aspe.hhs.gov/topics/poverty-economic-mobility/poverty-guidelines/prior-hhs-poverty-guidelines-federal-register-references/2017-poverty-guidelines).^3^

**eTable 2.** **Associations of adverse prenatal exposure (APE) burden with CBCL total problems at each time point**. Linear mixed-effect models were used to examine group difference in CBCL total problems among APE groups at each time point, adjusting for age, sex, pubertal stage, socioeconomic status, and random effects for subject ID, family ID, and site effects. All P value were FDR-corrected for five comparisons across time points.

| **Measures** | **N** | **Beta (SE)** | **T value** | ***P*_FDR_** | **Statistics** | ***P*_FDR_ overall** |
| --- | --- | --- | --- | --- | --- | --- |
| **Baseline** | | | | |  |  |
| One APE | 2039 | 0.099 (0.033) | 2.98 | 4.75E-03 |  |  |
| Two APEs | 1479 | 0.315 (0.036) | 8.74 | 1.46E-17 |  |  |
| Three+ APEs | 1083 | 0.625 (0.04) | 15.65 | 1.90E-53 | F = 98.57 | 1.48E-61 |
| **Year 1** | | | | |  |  |
| One APE | 2152 | 0.135 (0.032) | 4.16 | 1.64E-04 |  |  |
| Two APEs | 1524 | 0.27 (0.035) | 7.64 | 6.40E-14 |  |  |
| Three+ APEs | 1115 | 0.564 (0.04) | 14.27 | 4.57E-45 | F = 74.32 | 8.58E-47 |
| **Year 2** | |  |  |  |  |  |
| One APE | 2171 | 0.087 (0.033) | 2.65 | 1.00E-02 |  |  |
| Two APEs | 1590 | 0.226 (0.036) | 6.34 | 4.05E-10 |  |  |
| Three+ APEs | 1102 | 0.495 (0.04) | 12.43 | 8.17E-35 | F = 59.04 | 2.07E-37 |
| **Year 3** | |  |  |  |  |  |
| One APE | 2035 | 0.082 (0.034) | 2.42 | 1.57E-02 |  |  |
| Two APEs | 1477 | 0.222 (0.037) | 6.04 | 2.07E-09 |  |  |
| Three+ APEs | 1009 | 0.505 (0.042) | 12.15 | 1.79E-33 | F = 56.60 | 5.91E-36 |
| **Year 4** | |  |  |  |  |  |
| One APE | 989 | 0.155 (0.049) | 3.17 | 3.82E-03 |  |  |
| Two APEs | 674 | 0.283 (0.054) | 5.26 | 1.54E-07 |  |  |
| Three+ APEs | 438 | 0.564 (0.061) | 9.2 | 6.65E-20 | F = 30.29 | 3.07E-19 |

**eTable 3. Associations of adverse prenatal exposures (APE) burden with the risk of clinically significant psychopathology.** To assess cumulative effects of APEs on the risk of 17 clinically significant psychopathology, defined as CBCL composite scores ≥ 60 or sub-syndrome scores ≥ 65, we used generalized linear mixed-effects models with binomial error structure and logit link function *glmer* in R, adjusting for age, sex, pubertal stage, socioeconomic status, and random effects for subject ID, family ID, and site. Odds ratio (OR) and 95% confidence Interval (CI) were reported, with FDR corrected P value for 17 multiple corrections.

| **Measures** | **N** | **OR (95% CI)** | ***P*_FDR_** | **Statistics** | ***P*_FDR_ overall** |
| --- | --- | --- | --- | --- | --- |
| **Anxious/depressed** | | | |  |  |
| One APE | 2,565 | 1.34 (0.87-2.06) | 2.64E-01 |  |  |
| Two APEs | 1,871 | 2.05 (1.31-3.22) | 2.63E-03 |  |  |
| Three+ APEs | 1,349 | 2.14 (1.31-3.49) | 2.25E-03 | **Χ^2^ = 14.97** | **2.09E-03** |
| **Withdrawn/depressed** | | | |  |  |
| One APE | 2,565 | 1.19 (0.8-1.75) | 4.67E-01 |  |  |
| Two APEs | 1,871 | 1.48 (0.98-2.23) | 8.83E-02 |  |  |
| Three+ APEs | 1,349 | 1.99 (1.27-3.13) | 2.63E-03 | **Χ^2^ = 11.04** | **1.22E-02** |
| **Somatic complaints** | |  |  |  |  |
| One APE | 2,565 | 1.86 (1.28-2.7) | 2.41E-03 |  |  |
| Two APEs | 1,871 | 2.92 (2.01-4.23) | 1.44E-07 |  |  |
| Three+ APEs | 1,349 | 6.82 (4.52-10.29) | 5.33E-20 | **Χ^2^ = 98.46** | **1.89E-20** |
| **Social problems** | |  |  |  |  |
| One APE | 2,565 | 1.68 (0.8-3.54) | 2.64E-01 |  |  |
| Two APEs | 1,871 | 2.25 (1.05-4.83) | 5.40E-02 |  |  |
| Three+ APEs | 1,349 | 4.95 (2.35-10.43) | 4.59E-05 | **Χ^2^ = 23.43** | **5.15E-05** |
| **Thought problems** | |  |  |  |  |
| One APE | 2,565 | 0.78 (0.53-1.15) | 2.80E-01 |  |  |
| Two APEs | 1,871 | 0.79 (0.51-1.21) | 2.72E-01 |  |  |
| Three+ APEs | 1,349 | 1.21 (0.77-1.9) | 4.52E-01 | Χ^2^ = 5.55 | 1.35E-01 |
| **Attention problems** | |  |  |  |  |
| One APE | 2,565 | 1.7 (0.94-3.06) | 1.37E-01 |  |  |
| Two APEs | 1,871 | 2.69 (1.49-4.85) | 2.25E-03 |  |  |
| Three+ APEs | 1,349 | 4.95 (2.7-9.09) | 3.15E-07 | **Χ^2^ = 34.42** | **3.04E-07** |
| **Rule-breaking behaviors** | |  |  |  |  |
| One APE | 2,565 | 0.75 (0.75-0.75) | <0.001 |  |  |
| Two APEs | 1,871 | 1.43 (1.43-1.43) | <0.001 |  |  |
| Three+ APEs | 1,349 | 2.41 (2.41-2.41) | <0.001 | **Χ^2^ > 100** | **<0.001** |
| **Aggressive behaviors** | |  |  |  |  |
| One APE | 2,565 | 0.31 (0.17-0.58) | 6.43E-04 |  |  |
| Two APEs | 1,871 | 0.63 (0.36-1.11) | 1.27E-01 |  |  |
| Three+ APEs | 1,349 | 0.99 (0.56-1.75) | 9.77E-01 | **Χ^2^ = 18.16** | **5.32E-04** |
| **Internalizing problems** | |  |  |  |  |
| One APE | 2,565 | 1.38 (1.05-1.81) | 3.36E-02 |  |  |
| Two APEs | 1,871 | 2.34 (1.74-3.14) | 5.69E-08 |  |  |
| Three+ APEs | 1,349 | 5.26 (3.77-7.34) | 1.22E-22 | **Χ^2^ > 100** | **3.43E-24** |
| **Externalizing problems** | |  |  |  |  |
| One APE | 2,565 | 2.8 (1.72-4.57) | 1.82E-04 |  |  |
| Two APEs | 1,871 | 1.99 (1.17-3.38) | 1.77E-02 |  |  |
| Three+ APEs | 1,349 | 5.21 (3.07-8.84) | 2.78E-09 | **Χ^2^ = 40.44** | **1.83E-08** |
| **Total problems** | |  |  |  |  |
| One APE | 2,565 | 2.01 (1.28-3.16) | 6.46E-03 |  |  |
| Two APEs | 1,871 | 3.82 (2.39-6.11) | 9.12E-08 |  |  |
| Three+ APEs | 1,349 | 6.75 (4.14-11.02) | 1.31E-13 | **Χ^2^ = 69.72** | **1.67E-14** |
| **Depressive symptoms** | |  |  |  |  |
| One APE | 2,565 | 2.36 (1.6-3.5) | 1.50E-04 |  |  |
| Two APEs | 1,871 | 2.1 (1.39-3.16) | 1.21E-03 |  |  |
| Three+ APEs | 1,349 | 4.57 (2.97-7.04) | 1.97E-11 | **Χ^2^ = 47.92** | **6.26E-10** |
| **Anxiety symptoms** | |  |  |  |  |
| One APE | 2,565 | 1.03 (0.68-1.56) | 8.82E-01 |  |  |
| Two APEs | 1,871 | 1.28 (0.83-1.98) | 2.70E-01 |  |  |
| Three+ APEs | 1,349 | 2.25 (1.43-3.53) | 5.28E-04 | **Χ^2^ = 17.48** | **6.84E-04** |
| **Somatic symptoms** | |  |  |  |  |
| One APE | 2,565 | 1.4 (1.09-1.81) | 2.34E-02 |  |  |
| Two APEs | 1,871 | 2.1 (1.59-2.76) | 7.04E-07 |  |  |
| Three APEs | 1,349 | 3.6 (2.68-4.83) | 5.74E-16 | **Χ^2^ = 78.81** | **2.35E-16** |
| **ADHD symptoms** | |  |  |  |  |
| One APE | 2,565 | 5 (2.33-10.74) | 1.68E-04 |  |  |
| Two APEs | 1,871 | 6.82 (3.18-14.65) | 2.77E-06 |  |  |
| Three+ APEs | 1,349 | 11.36 (5.19-24.88) | 1.87E-09 | **Χ^2^ = 40.5** | **1.83E-08** |
| **Oppositional defiant symptoms** | |  |  |  |  |
| One APE | 2,565 | 1.23 (0.69-2.22) | 5.23E-01 |  |  |
| Two APEs | 1,871 | 1.72 (0.93-3.15) | 9.76E-02 |  |  |
| Three+ APEs | 1,349 | 3.22 (1.75-5.92) | 2.12E-04 | **Χ^2^ = 19.58** | **2.95E-04** |
| **Conduct symptoms** | |  |  |  |  |
| One APE | 2,565 | 1.43 (0.69-2.96) | 3.98E-01 |  |  |
| Two APEs | 1,871 | 2.16 (1.05-4.46) | 5.40E-02 |  |  |
| Three+ APEs | 1,349 | 4.35 (2.15-8.81) | 8.28E-05 | **Χ^2^ = 23.4** | **5.15E-05** |

Abbreviations: ADHD, attention-deficit/hyperactivity disorder.

**eTable 4. Associations between adverse prenatal exposure (APE) burden and CBCL scores.** We tested associations between APEs and 17 CBCL scores using linear mixed-effects models, adjusting for age, sex, pubertal stage, socioeconomic status, and random effects for subject ID, family ID, and site. Within each model, CBCL scores were treated as continuous dependent variables, with cumulative APE burden as an independent predictor. Standardized beta coefficients and standard errors (SE) were reported for interaction effects between age and each APE level, and F statistics were used for overall interaction effects, with FDR corrected *P* value for 17 comparisons.

| **Measures** | **N** | **Beta (SE)** | **T value** | ***P*_FDR_** | **Statistics** | ***P*_FDR_ overall** |
| --- | --- | --- | --- | --- | --- | --- |
| **Anxious/depressed** | | | | |  |  |
| One APE | 2,565 | 0.065 (0.027) | 2.43 | 1.70E-02 |  |  |
| Two APEs | 1,871 | 0.131 (0.029) | 4.53 | 5.91E-06 |  |  |
| Three+ APEs | 1,349 | 0.351 (0.032) | 10.91 | 1.70E-27 | F = 44.453 | 1.97E-28 |
| **Withdrawn/depressed** | | | | |  |  |
| One APE | 2,565 | 0.072 (0.025) | 2.83 | 5.74E-03 |  |  |
| Two APEs | 1,871 | 0.143 (0.028) | 5.18 | 2.65E-07 |  |  |
| Three+ APEs | 1,349 | 0.32 (0.031) | 10.46 | 2.01E-25 | F = 40.066 | 1.13E-25 |
| **Somatic complaints** | |  |  |  |  |  |
| One APE | 2,565 | 0.086 (0.025) | 3.48 | 1.30E-03 |  |  |
| Two APEs | 1,871 | 0.216 (0.027) | 8 | 8.50E-15 |  |  |
| Three+ APEs | 1,349 | 0.402 (0.03) | 13.35 | 4.83E-40 | F = 68.662 | 1.42E-43 |
| **Social problems** | |  |  |  |  |  |
| One APE | 2,565 | 0.078 (0.026) | 2.96 | 4.38E-03 |  |  |
| Two APEs | 1,871 | 0.213 (0.029) | 7.4 | 3.23E-13 |  |  |
| Three+ APEs | 1,349 | 0.472 (0.032) | 14.81 | 2.79E-48 | F = 85.198 | 1.56E-53 |
| **Thought problems** | |  |  |  |  |  |
| One APE | 2,565 | 0.076 (0.026) | 2.88 | 5.28E-03 |  |  |
| Two APEs | 1,871 | 0.204 (0.029) | 7.14 | 1.80E-12 |  |  |
| Three+ APEs | 1,349 | 0.463 (0.032) | 14.62 | 2.55E-47 | F = 82.859 | 2.95E-52 |
| **Attention problems** | |  |  |  |  |  |
| One APE | 2,565 | 0.091 (0.027) | 3.31 | 1.61E-03 |  |  |
| Two APEs | 1,871 | 0.233 (0.03) | 7.86 | 1.86E-14 |  |  |
| Three+ APEs | 1,349 | 0.484 (0.033) | 14.73 | 6.90E-48 | F = 83.805 | 8.84E-53 |
| **Rule-breaking behaviors** | |  |  |  |  |  |
| One APE | 2,565 | 0.081 (0.026) | 3.12 | 2.78E-03 |  |  |
| Two APEs | 1,871 | 0.22 (0.028) | 7.83 | 1.89E-14 |  |  |
| Three+ APEs | 1,349 | 0.51 (0.031) | 16.39 | 2.45E-58 | F = 104.474 | 2.83E-65 |
| **Aggressive behaviors** | |  |  |  |  |  |
| One APE | 2,565 | 0.091 (0.027) | 3.36 | 1.47E-03 |  |  |
| Two APEs | 1,871 | 0.202 (0.029) | 6.84 | 1.35E-11 |  |  |
| Three+ APEs | 1,349 | 0.46 (0.033) | 14.08 | 3.82E-44 | F = 74.175 | 5.59E-47 |
| **Internalizing problems** | |  |  |  |  |  |
| One APE | 2,565 | 0.09 (0.026) | 3.4 | 1.41E-03 |  |  |
| Two APEs | 1,871 | 0.192 (0.029) | 6.7 | 3.09E-11 |  |  |
| Three+ APEs | 1,349 | 0.432 (0.032) | 13.5 | 7.05E-41 | F = 67.55 | 6.49E-43 |
| **Externalizing problems** | |  |  |  |  |  |
| One APE | 2,565 | 0.094 (0.027) | 3.46 | 1.30E-03 |  |  |
| Two APEs | 1,871 | 0.221 (0.029) | 7.52 | 1.53E-13 |  |  |
| Three+ APEs | 1,349 | 0.507 (0.033) | 15.55 | 6.35E-53 | F = 91.666 | 1.78E-57 |
| **Total problems** | |  |  |  |  |  |
| One APE | 2,565 | 0.108 (0.027) | 4.02 | 6.46E-04 |  |  |
| Two APEs | 1,871 | 0.261 (0.029) | 8.9 | 1.23E-17 |  |  |
| Three+ APEs | 1,349 | 0.576 (0.033) | 17.63 | 6.68E-67 | F = 117.644 | 3.98E-73 |
| **Depressive symptoms** | |  |  |  |  |  |
| One APE | 2,565 | 0.097 (0.026) | 3.78 | 6.72E-04 |  |  |
| Two APEs | 1,871 | 0.172 (0.028) | 6.19 | 7.88E-10 |  |  |
| Three+ APEs | 1,349 | 0.388 (0.031) | 12.52 | 1.65E-35 | F = 56.106 | 9.18E-36 |
| **Anxiety symptoms** | |  |  |  |  |  |
| One APE | 2,565 | 0.06 (0.027) | 2.27 | 2.34E-02 |  |  |
| Two APEs | 1,871 | 0.147 (0.029) | 5.08 | 4.20E-07 |  |  |
| Three+ APEs | 1,349 | 0.373 (0.032) | 11.63 | 6.34E-31 | F = 51.781 | 4.71E-33 |
| **Somatic symptoms** | |  |  |  |  |  |
| One APE | 2,565 | 0.09 (0.024) | 3.68 | 7.96E-04 |  |  |
| Two APEs | 1,871 | 0.206 (0.026) | 7.79 | 2.14E-14 |  |  |
| Three APEs | 1,349 | 0.37 (0.029) | 12.55 | 1.24E-35 | F = 59.912 | 3.95E-38 |
| **ADHD symptoms** | |  |  |  |  |  |
| One APE | 2,565 | 0.105 (0.027) | 3.86 | 6.46E-04 |  |  |
| Two APEs | 1,871 | 0.249 (0.03) | 8.44 | 3.17E-16 |  |  |
| Three+ APEs | 1,349 | 0.48 (0.033) | 14.66 | 1.46E-47 | F = 81.918 | 9.62E-52 |
| **Oppositional defiant symptoms** | |  |  |  |  |  |
| One APE | 2,565 | 0.106 (0.027) | 3.93 | 6.46E-04 |  |  |
| Two APEs | 1,871 | 0.21 (0.029) | 7.2 | 1.23E-12 |  |  |
| Three+ APEs | 1,349 | 0.443 (0.032) | 13.73 | 3.89E-42 | F = 68.929 | 1.01E-43 |
| **Conduct symptoms** | |  |  |  |  |  |
| One APE | 2,565 | 0.061 (0.027) | 2.3 | 2.28E-02 |  |  |
| Two APEs | 1,871 | 0.193 (0.029) | 6.67 | 3.70E-11 |  |  |
| Three+ APEs | 1,349 | 0.454 (0.032) | 14.16 | 1.28E-44 | F = 80.028 | 1.35E-50 |

Abbreviations: ADHD, attention-deficit/hyperactivity disorder.

**eTable 5. Interaction effects of adverse prenatal exposure (APE) burden and age on CBCL developmental trajectories.** To examine APE-by-age interactions on CBCL developmental trajectories, linear mixed-effects models were conducted, where CBCL scores were modeled as continuous dependent variables, with APE group, age, and APE-by-age interaction term as fixed effects, adjusting for sex, pubertal stage, socioeconomic status, and random effects for subject ID, family ID, and site. Standardized beta coefficients and standard errors (SE) were reported for interaction effects between age and each APE level, and F statistics were used for overall interaction effects, with FDR corrected *P* value for 17 comparisons.

| **Measures** | **N** | **Beta (SE)** | **T value** | ***P*_FDR_** | **Statistics** | ***P*_FDR_ overall** |
| --- | --- | --- | --- | --- | --- | --- |
| **Anxious/depressed** | | | | |  |  |
| One APE | 2,565 | 0.007 (0.011) | 0.64 | 9.59E-01 |  |  |
| Two APEs | 1,871 | -0.023 (0.012) | -1.9 | 8.81E-02 |  |  |
| Three+ APEs | 1,349 | -0.016 (0.013) | -1.21 | 3.19E-01 | **F = 3.01** | **4.47E-02** |
| **Withdrawn/depressed** | | | | |  |  |
| One APE | 2,565 | 0.004 (0.012) | 0.3 | 9.59E-01 |  |  |
| Two APEs | 1,871 | 0.012 (0.013) | 0.92 | 4.35E-01 |  |  |
| Three+ APEs | 1,349 | 0.032 (0.015) | 2.23 | 4.35E-02 | F = 1.99 | 1.60E-01 |
| **Somatic complaints** | |  |  |  |  |  |
| One APE | 2,565 | -0.018 (0.013) | -1.45 | 9.59E-01 |  |  |
| Two APEs | 1,871 | -0.03 (0.014) | -2.22 | 6.44E-02 |  |  |
| Three+ APEs | 1,349 | -0.006 (0.015) | -0.38 | 8.01E-01 | F = 1.92 | 1.62E-01 |
| **Social problems** | |  |  |  |  |  |
| One APE | 2,565 | -0.002 (0.011) | -0.14 | 9.59E-01 |  |  |
| Two APEs | 1,871 | -0.051 (0.012) | -4.21 | 4.30E-04 |  |  |
| Three+ APEs | 1,349 | -0.063 (0.013) | -4.72 | 2.01E-05 | **F = 14.24** | **4.88E-08** |
| **Thought problems** | |  |  |  |  |  |
| One APE | 2,565 | -0.006 (0.011) | -0.49 | 9.59E-01 |  |  |
| Two APEs | 1,871 | -0.039 (0.012) | -3.14 | 1.43E-02 |  |  |
| Three+ APEs | 1,349 | -0.028 (0.014) | -2.06 | 6.09E-02 | **F = 4.56** | **7.14E-03** |
| **Attention problems** | |  |  |  |  |  |
| One APE | 2,565 | -0.003 (0.01) | -0.32 | 9.59E-01 |  |  |
| Two APEs | 1,871 | -0.021 (0.011) | -1.94 | 8.81E-02 |  |  |
| Three+ APEs | 1,349 | -0.035 (0.012) | -3 | 7.59E-03 | **F = 4.22** | **1.04E-02** |
| **Rule-breaking behaviors** | |  |  |  |  |  |
| One APE | 2,565 | 0.014 (0.012) | 1.2 | 9.59E-01 |  |  |
| Two APEs | 1,871 | 0.004 (0.012) | 0.34 | 7.32E-01 |  |  |
| Three+ APEs | 1,349 | 0.015 (0.014) | 1.06 | 3.77E-01 | F = 0.67 | 5.71E-01 |
| **Aggressive behaviors** | |  |  |  |  |  |
| One APE | 2,565 | -0.001 (0.01) | -0.09 | 9.59E-01 |  |  |
| Two APEs | 1,871 | -0.023 (0.011) | -2.07 | 8.12E-02 |  |  |
| Three+ APEs | 1,349 | -0.046 (0.012) | -3.69 | 9.39E-04 | **F = 6.59** | **8.03E-04** |
| **Internalizing problems** | |  |  |  |  |  |
| One APE | 2,565 | -0.001 (0.011) | -0.09 | 9.59E-01 |  |  |
| Two APEs | 1,871 | -0.018 (0.012) | -1.47 | 2.00E-01 |  |  |
| Three+ APEs | 1,349 | 0.001 (0.013) | 0.06 | 9.50E-01 | F = 1.09 | 3.83E-01 |
| **Externalizing problems** | |  |  |  |  |  |
| One APE | 2,565 | 0.004 (0.01) | 0.42 | 9.59E-01 |  |  |
| Two APEs | 1,871 | -0.015 (0.011) | -1.39 | 2.17E-01 |  |  |
| Three+ APEs | 1,349 | -0.029 (0.012) | -2.33 | 4.27E-02 | **F = 3.43** | **2.77E-02** |
| **Total problems** | |  |  |  |  |  |
| One APE | 2,565 | -0.001 (0.01) | -0.05 | 9.59E-01 |  |  |
| Two APEs | 1,871 | -0.031 (0.011) | -2.91 | 1.64E-02 |  |  |
| Three+ APEs | 1,349 | -0.031 (0.012) | -2.58 | 2.43E-02 | **F = 5.51** | **2.34E-03** |
| **Depressive symptoms** | |  |  |  |  |  |
| One APE | 2,565 | -0.001 (0.012) | -0.08 | 9.59E-01 |  |  |
| Two APEs | 1,871 | 0.005 (0.013) | 0.4 | 7.32E-01 |  |  |
| Three+ APEs | 1,349 | 0.051 (0.014) | 3.54 | 1.37E-03 | **F = 5.82** | **1.94E-03** |
| **Anxiety symptoms** | |  |  |  |  |  |
| One APE | 2,565 | 0.005 (0.011) | 0.47 | 9.59E-01 |  |  |
| Two APEs | 1,871 | -0.032 (0.012) | -2.62 | 2.98E-02 |  |  |
| Three+ APEs | 1,349 | -0.03 (0.014) | -2.24 | 4.35E-02 | **F = 5.45** | **2.34E-03** |
| **Somatic symptoms** | |  |  |  |  |  |
| One APE | 2,565 | -0.021 (0.013) | -1.58 | 9.59E-01 |  |  |
| Two APEs | 1,871 | -0.033 (0.014) | -2.3 | 6.12E-02 |  |  |
| Three APEs | 1,349 | -0.014 (0.016) | -0.89 | 4.53E-01 | F = 1.85 | 1.65E-01 |
| **ADHD symptoms** | |  |  |  |  |  |
| One APE | 2,565 | -0.008 (0.01) | -0.84 | 9.59E-01 |  |  |
| Two APEs | 1,871 | -0.031 (0.011) | -2.89 | 1.64E-02 |  |  |
| Three+ APEs | 1,349 | -0.067 (0.012) | -5.75 | 1.56E-07 | **F = 13.51** | **7.13E-08** |
| **Oppositional defiant symptoms** | |  |  |  |  |  |
| One APE | 2,565 | -0.003 (0.011) | -0.24 | 9.59E-01 |  |  |
| Two APEs | 1,871 | -0.022 (0.012) | -1.92 | 8.81E-02 |  |  |
| Three+ APEs | 1,349 | -0.055 (0.013) | -4.24 | 1.28E-04 | **F = 7.88** | **1.69E-04** |
| **Conduct symptoms** | |  |  |  |  |  |
| One APE | 2,565 | 0.012 (0.011) | 1.06 | 9.59E-01 |  |  |
| Two APEs | 1,871 | -0.007 (0.012) | -0.61 | 6.15E-01 |  |  |
| Three+ APEs | 1,349 | 0.001 (0.013) | 0.1 | 9.50E-01 | F = 1.07 | 3.83E-01 |

Abbreviations: ADHD, attention-deficit/hyperactivity disorder.

**eTable 15. Baseline characteristics of sibling pairs with discordant adverse prenatal exposures (APEs)**

|  | **Participants, No. (%)** | | |  |  |
| --- | --- | --- | --- | --- | --- |
| **Characteristic** | **Total**  **(n =** **828)** | **No APE**  **(n = 414)** | **One APE**  **(n = 414)** | **Statistics** | ***P* value** |
| **Female, No. (%)** | 423 (51.1%) | 216 (52.2%) | 207 (50%) | Χ^2^ = 0.37 | 9.46E-01 |
| **Age, y** | 9.8 (0.7) | 9.8 (0.7) | 9.9 (0.7) | F = 3.23 | 2.18E-02 |
| **Pubertal stage** | 1.9 (0.7) | 1.8 (0.7) | 1.9 (0.8) | F = 4.3 | 5.13E-03 |
| **Intracranial volume** | 1,506,944.1 (159,655) | 1,509,378.4 (157,374) | 1,504,515.9 (162,056.7) | F = 4.13 | 6.44E-03 |
| **Surface hole number** | 26.9 (12.3) | 26.9 (12.3) | 26.9 (12.2) | F = 0.74 | 5.31E-01 |
| **Scanner** |  |  |  |  |  |
| General Electric | 478 (64.3) | 245 (64.8) | 233 (63.8) |  |  |
| Philips | 53 (7.1) | 30 (7.9) | 23 (6.3) |  |  |
| Siemens | 212 (28.5) | 103 (27.2) | 109 (29.9) | Χ^2^ =8 | 2.38E-01 |
| **Prenatal exposures** |  |  |  |  |  |
| Unplanned pregnancy | 341 (41.2%) | 105 (25.4%) | 236 (57%) | Χ^2^ = 244.29 | 1.12E-52 |
| Early alcohol exposure | 160 (19.3%) | 42 (10.1%) | 118 (28.5%) | Χ^2^ = 178.39 | 1.96E-38 |
| Early tobacco exposure | 73 (8.8%) | 22 (5.3%) | 51 (12.3%) | Χ^2^ = 111.7 | 4.74E-24 |
| Early marijuana exposure | 21 (2.5%) | 4 (1%) | 17 (4.1%) | Χ^2^ = 73.05 | 9.46E-16 |
| Complicated pregnancy | 320 (38.6%) | 83 (20%) | 237 (57.2%) | Χ^2^ = 253.38 | 1.21E-54 |
| Complicated birth | 206 (24.9%) | 34 (8.2%) | 172 (41.5%) | Χ^2^ = 172.69 | 3.34E-37 |
| **Child Behavior Checklist (CBCL) measures at baseline** | | | |  |  |
| Anxious/depressed | 53.4 (5.8) | 53.1 (5.5) | 53.6 (6.0) | F = 1.23 | 2.99E-01 |
| Withdrawn/depressed | 53.5 (5.9) | 53.4 (5.6) | 53.5 (6.2) | F = 1.79 | 1.48E-01 |
| Somatic complaints | 54.3 (5.7) | 54.4 (5.8) | 54.2 (5.6) | F = 3.62 | 1.29E-02 |
| Social problems | 52.6 (4.5) | 52.4 (4.0) | 52.8 (4.9) | F = 5.61 | 8.28E-04 |
| Thought problems | 53.4 (5.7) | 53.2 (5.7) | 53.6 (5.8) | F = 0.93 | 4.24E-01 |
| Attention problems | 53.2 (5.5) | 52.8 (4.7) | 53.5 (6.1) | F = 6.36 | 2.89E-04 |
| Rule-breaking behaviors | 52.7 (4.8) | 52.4 (4.7) | 52.9 (5.0) | F = 12.12 | 9.05E-08 |
| Aggressive behaviors | 53.0 (5.5) | 52.8 (5.3) | 53.2 (5.6) | F = 7.07 | 1.08E-04 |
| Internalizing problems | 47.9 (10.4) | 47.8 (10.3) | 48.1 (10.6) | F = 2.64 | 4.85E-02 |
| Externalizing problems | 45.7 (10.5) | 45.2 (10.4) | 46.2 (10.7) | F = 8.99 | 7.30E-06 |
| Total problems | 45.2 (11.2) | 44.6 (11.1) | 45.7 (11.3) | F = 8.34 | 1.80E-05 |
| Depressive symptoms | 53.3 (5.4) | 53.1 (5.2) | 53.5 (5.7) | F = 2.92 | 3.31E-02 |
| Anxiety symptoms | 53.1 (5.6) | 52.9 (5.7) | 53.2 (5.6) | F = 1.16 | 3.26E-01 |
| Somatic symptoms | 54.9 (6.2) | 55.0 (6.4) | 54.7 (6.1) | F = 2.36 | 7.00E-02 |
| ADHD symptoms | 52.6 (4.8) | 52.3 (4.4) | 52.8 (5.1) | F = 4.79 | 2.59E-03 |
| Oppositional defiant symptoms | 53.5 (5.5) | 53.3 (5.3) | 53.7 (5.7) | F = 7.02 | 1.16E-04 |
| Conduct symptoms | 53.2 (5.8) | 52.9 (5.6) | 53.6 (6) | F = 11.45 | 2.34E-07 |

Abbreviations: ADHD, attention-deficit/hyperactivity disorder. Categorical variables were assessed using Chi-square tests, and quantitative measures using analysis of variance (ANOVA) among adverse prenatal exposure groups. Quantitative measures are presented as means and standard deviations (SDs); categorical variables are presented as counts and percentages.

**eTable 16. Associations of adverse prenatal exposure (APE) group with CBCL scores in sibling pairs.** We tested associations between APE group and 17 CBCL scores in sibling pairs using linear mixed-effects models, adjusting for age, sex, pubertal stage, socioeconomic status, and random effects for subject ID, family ID, and site. Within each model, CBCL scores were treated as continuous dependent variables, with APE group in sibling pairs as an independent predictor. Standardized beta coefficients and standard errors (SE) for group differences between more-exposed and less-exposed groups were reported, with uncorrected *P* value.

|  | **More- vs. less-exposed siblings** | | |
| --- | --- | --- | --- |
| **Measures** | **Beta (SE)** | **T value** | ***P* value** |
| Anxious/depressed | 0.054 (0.049) | 1.1 | 2.73E-01 |
| Withdrawn/depressed | 0.044 (0.052) | 0.86 | 3.93E-01 |
| Somatic complaints | 0.018 (0.047) | 0.38 | 7.03E-01 |
| Social problems | 0.091 (0.051) | 1.78 | 7.63E-02 |
| Thought problems | 0.101 (0.053) | 1.91 | 5.67E-02 |
| Attention problems | 0.064 (0.058) | 1.11 | 2.68E-01 |
| **Rule-breaking behaviors** | **0.133 (0.054)** | **2.48** | **1.38E-02** |
| **Aggressive behaviors** | **0.118 (0.054)** | **2.19** | **2.91E-02** |
| Internalizing problems | 0.051 (0.047) | 1.08 | 2.81E-01 |
| **Externalizing problems** | **0.13 (0.054)** | **2.41** | **1.67E-02** |
| **Total problems** | **0.111 (0.049)** | **2.25** | **2.48E-02** |
| **Depressive symptoms** | **0.134 (0.052)** | **2.59** | **1.01E-02** |
| Anxiety symptoms | 0.03 (0.051) | 0.6 | 5.50E-01 |
| Somatic symptoms | -0.014 (0.048) | -0.3 | 7.65E-01 |
| ADHD symptoms | 0.073 (0.056) | 1.3 | 1.94E-01 |
| Oppositional defiant symptoms | 0.101 (0.056) | 1.79 | 7.34E-02 |
| **Conduct symptoms** | **0.14 (0.055)** | **2.54** | **1.15E-02** |

**eTable 17. Interaction effects of adverse prenatal exposure (APE) and age on CBCL developmental trajectories in sibling pairs.** To examine APE-by-age interactions on CBCL developmental trajectories in sibling pairs, linear mixed-effects models were conducted, where CBCL scores were modeled as continuous dependent variables, with APE group, age, and APE-by-age interaction term as fixed effects, adjusting for sex, pubertal stage, socioeconomic status, and random effects for subject ID, family ID, and site. Standardized beta coefficients and standard errors (SE) were reported for interaction effects between age and more-exposed group compared to less-exposed group, with uncorrected *P* value.

|  | **More- vs. less-exposed siblings** | | |
| --- | --- | --- | --- |
| **Measures** | **Beta (SE)** | **T value** | ***P* value** |
| Anxious/depressed | -0.018 (0.028) | -0.63 | 5.30E-01 |
| Withdrawn/depressed | 0.014 (0.028) | 0.52 | 6.06E-01 |
| Somatic complaints | 0.005 (0.028) | 0.19 | 8.47E-01 |
| Social problems | -0.018 (0.025) | -0.72 | 4.75E-01 |
| Thought problems | -0.012 (0.026) | -0.48 | 6.32E-01 |
| Attention problems | -0.017 (0.025) | -0.69 | 4.91E-01 |
| Rule-breaking behaviors | -0.013 (0.025) | -0.54 | 5.90E-01 |
| Aggressive behaviors | 0.031 (0.029) | 1.06 | 2.87E-01 |
| Internalizing problems | -0.028 (0.028) | -0.99 | 3.24E-01 |
| Externalizing problems | -0.002 (0.025) | -0.06 | 9.49E-01 |
| Total problems | -0.014 (0.026) | -0.53 | 5.97E-01 |
| Depressive symptoms | -0.018 (0.028) | -0.63 | 5.30E-01 |
| Anxiety symptoms | 0.014 (0.028) | 0.52 | 6.06E-01 |
| Somatic symptoms | 0.005 (0.028) | 0.19 | 8.47E-01 |
| ADHD symptoms | -0.018 (0.025) | -0.72 | 4.75E-01 |
| Oppositional defiant symptoms | -0.012 (0.026) | -0.48 | 6.32E-01 |
| Conduct symptoms | -0.017 (0.025) | -0.69 | 4.91E-01 |
